# Information bias of vaccine effectiveness estimation due to informed consent for national registration of COVID-19 vaccination

**DOI:** 10.1101/2023.05.23.23290384

**Authors:** C.H. (Henri) van Werkhoven, Brechje de Gier, Scott McDonald, Hester E. de Melker, Susan J.M. Hahné, Susan van den Hof, Mirjam J. Knol

## Abstract

**Background:** Registration in the Dutch national COVID-19 vaccination register requires consent from the vaccinee. This causes misclassification of non-consenting vaccinated persons as being unvaccinated. We quantified and corrected the resulting information bias in the estimation of vaccine effectiveness (VE).

**Methods:** National data were used for the period dominated by the SARS-CoV-2 Delta variant (11 July to 15 November 2021). VE ((1-relative risk)*100 %) against COVID-19 hospitalization and ICU admission was estimated for individuals 12-49, 50-69, and ≥70 years of age using negative binomial regression. Anonymous data on vaccinations administered by the Municipal Health Services were used to determine informed consent percentages and estimate corrected VEs by iteratively imputing corrected vaccination status. Absolute bias was calculated as the absolute change in VE; relative bias as uncorrected / corrected relative risk.

**Results:** A total of 8,804 COVID-19 hospitalizations and 1,692 COVID-19 ICU admissions were observed. The bias was largest in the 70+ age group where the non-consent proportion was 7.0% and observed vaccination coverage was 87%: VE of primary vaccination against hospitalization changed from 75.5% (95% CI 73.5-77.4) before to 85.9% (95% CI 84.7-87.1) after correction (absolute bias -10.4 percentage point, relative bias 1.74). VE against ICU admission in this group was 88.7% (95% CI 86.2-90.8) before and 93.7% (95% CI 92.2-94.9) after correction (absolute bias -5.0 percentage point, relative bias 1.79).

**Conclusions:** VE estimates can be substantially biased with modest non-consent percentages for registration of vaccination. Data on covariate specific non-consent percentages should be available to correct this bias.

## INTRODUCTION

Monitoring vaccine coverage and vaccine effectiveness (VE) is important to support infectious disease control and vaccine policy making.(1) Analyzing data from nationwide COVID-19 vaccination registries linked to outcome data such as hospitalizations is essential to provide rapid evidence on VE to inform timely public health decisions to optimally control the COVID-19 pandemic.

In the Netherlands, individual informed consent has to be obtained to share vaccination status in national vaccination register at the National Institute of Public Health, while vaccination information is available in the medical records that are stored at the vaccination provider (e.g. the municipal health service, general practitioner or nursing home). To the best of our knowledge, this strict interpretation of the General Data Protection Regulation (GDPR) is unique for the Netherlands. It is operationalized in the Dutch national COVID-vaccination Information and Monitoring System (CIMS), which includes only those vaccination records of individuals who provided consent for registration in CIMS; it does not include any data from the non-vaccinated or non-consenting vaccinated individuals. As a result, vaccine coverage data based on CIMS are incomplete. Moreover, linkage of this database to other data sources such as hospitalization records inevitably results in misclassification of vaccination status: vaccinated individuals without consent will be erroneously classified as unvaccinated due to the absence of a vaccination record in CIMS. Hence, while informed consent generally leads to a risk of selection bias with a generalizability concern, the way it was implemented here leads to potential information bias leading to concerns of internal validity.

All healthcare providers administering COVID-19 vaccines contribute to CIMS. In addition, anonymous data about the informed consent status is available from all individuals vaccinated by the Municipal Health Services (Dutch: *Gemeentelijke Gezondheidsdienst*, GGD). Since the GGDs administered 86% of COVID-19 primary series vaccinations in the Netherlands,(2) this data source can provide a good indication of the amount of misclassification in the population over time. Although these data cannot be linked to individual hospitalization records, the availability of this dataset provides a unique opportunity to quantify the information bias resulting from the informed consent requirement.

Since the summer of 2021, we have monitored the VE against hospitalization utilizing linkage of hospitalization to CIMS data.(3) In this study we analyzed to what extent non-consent may have biased the estimated VE against COVID-19 hospitalization and ICU admission in adults.

## Methods

### Bias estimation by formula

To get insight in the relative importance of different parameters, prior to modelling VE information bias present in real data, we calculated the bias in the absence of confounding factors and assuming a constant vaccination uptake and non-consent percentage. This was based on a formula with three parameters: the true percentage vaccinated in the population (*v*), the percentage of vaccinated persons not providing informed consent (*nc*), and the true relative risk (*RR*) of the outcome from which VE is calculated as (1-RR) * 100%. We used the following formula for the observed RR:

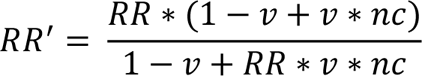

(see derivation in supplementary appendix). We visualized the relation between these parameters and the resulting relative bias (calculated as observed RR divided by true RR) and absolute bias (calculated as true VE minus observed VE in percentage points) by calculating the observed RR for each combination of parameters.

### Data sources

To quantify the information bias for the VE against COVID-19 hospitalization and ICU admission in the Netherlands we used three data sources, all stratified by year of birth, sex, region and calendar date, for the period 11 July to 15 November 2021. This period was dominated by the SARS-CoV-2 Delta variant.

1. Original population dataset: Dutch Personal Records Database for the total population size on January 1st, 2021, enriched with CIMS for daily vaccination uptake.
2. Non-consent percentages for COVID-19 vaccination based on anonymous data from the Municipal Health Services (GGDs).
3. Hospital admissions, from the Dutch National Intensive Care Evaluation (NICE) COVID-19 registry, which consists of all hospitalized persons with a positive SARS-CoV-2 test or Computed Tomography confirmed COVID-19.

For a full description of the data sources and justification of data selection and preparation, please refer to the supplementary methods.

### Vaccination status

Four COVID-19 vaccines were used in the Netherlands within the study period: Comirnaty (BNT162b2; BioNTech/Pfizer, Mainz, Germany/New York, United States (US)), Spikevax (mRNA-1273, Moderna, Cambridge, US), Vaxzevria (ChAdOx1-S; AstraZeneca, Cambridge, United Kingdom), and Jcovden (Ad26.COV2-S (recombinant), Janssen-Cilag International NV, Beerse, Belgium). The *primary series vaccination* included either one dose of Jcovden or two doses of any of the other vaccines. Vaccination status was categorized as unvaccinated (up to 14 days after receipt of the first vaccination dose), partly vaccinated (from 14 days after the first dose, up to either 28 days after the first dose in case of the Jcovden vaccine or 14 days after receipt of a second dose), or fully vaccinated (having received the Jcovden vaccine ≥ 28 days ago or a second dose of one of the other vaccines ≥ 14 days ago) in line with the analyses used in the Dutch national COVID-19 surveillance reports up to November 2021.(3, 4) We did not take the vaccine type or time since vaccination into account in our analyses, i.e. we assumed the VE to be the same for all vaccine types and to be stable over time.

### Data analysis

We used a negative binomial regression model to estimate the RR for partly and fully vaccinated individuals as compared to unvaccinated, using the original events and population datasets. VE was calculated as (1-RR) * 100%. Next, we performed two different imputation approaches to correct the VE. In the first, ‘Partially corrected analysis’, we assumed that individuals vaccinated by providers other than the GGD (e.g. general practitioners) had all provided informed consent. Consequently, the observed number of non-consenting individuals from the GGD were used for correction of the denominator data. In the second, ‘Fully corrected analysis’, we analyzed non-consent percentages by birth year, sex, region and calendar time, and generalized these to individuals vaccinated by providers other than the GGD, thus assuming that non-consent percentages were similar conditional on these covariates.

Consequently, the observed number of non-consenting individuals, incremented by the predicted number of non-consenting individuals from other providers, were used for correction of the denominator data. In both imputation approaches, the amount of misclassification in the numerator was predicted, assuming that the incidence of an outcome was independent of providing informed consent, conditional on the covariates in the imputation model. We repeated the negative binomial regression model to estimate the corrected VEs. Since the correction of the numerator was based on a biased estimate of the VE, the imputation process was repeated using the corrected VE until convergence was reached. Analyses were stratified for age groups 12-49 years, 50-69 years, and 70+ years old. Please refer to the supplementary methods for a full description of the statistical methods.

Analyses were performed in R version 4.1.2.(5) For regressions we used the mgcv package version 1.8-38.(6)

## Results

### Bias estimation by formula

Using the information bias formula, the relative bias of the true RR increased with increasing true VE, vaccination coverage, and non-consent percentage, whereas the absolute bias of the VE increased with increasing vaccination coverage and non-consent percentage, and was maximal at a true VE of around 50-60% (Figure 2A, 2B). Keeping vaccination coverage and non-consent percentage constant, a decline in VE (e.g. due to waning of the VE over time) led to a slight increase in the bias over time (Figure 2C).

**Figure 1:**
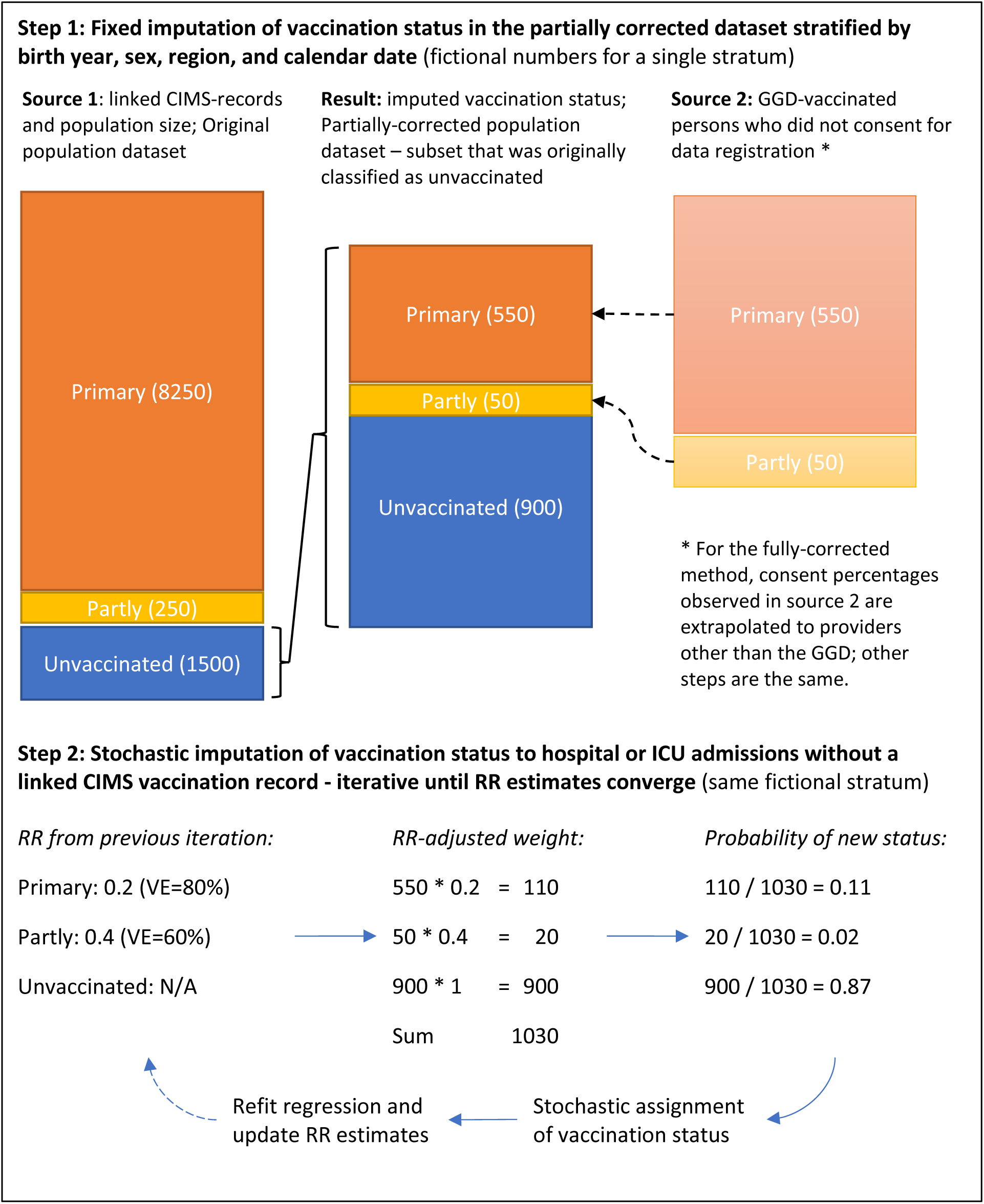
Method for correction of vaccination status. Fixed imputation of vaccination status in the population dataset followed by stochastic imputation of vaccination status for hospitalized patients without a linked CIMS vaccination record. Abbreviations: CIMS: Corona Information and Monitoring System. GGD: Municipal Health Services. RR: relative risk. N/A: not applicable.

**Figure 2:**
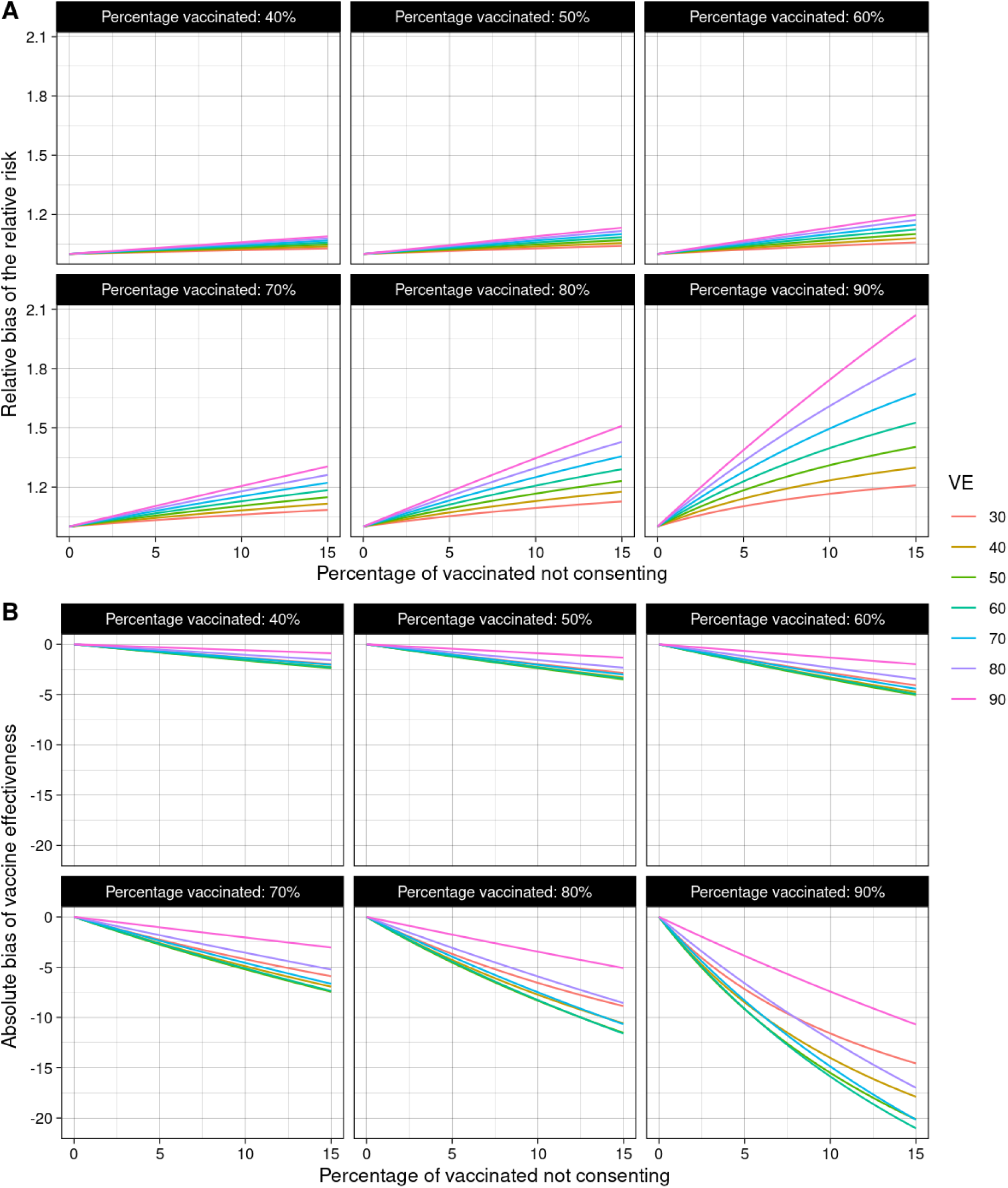
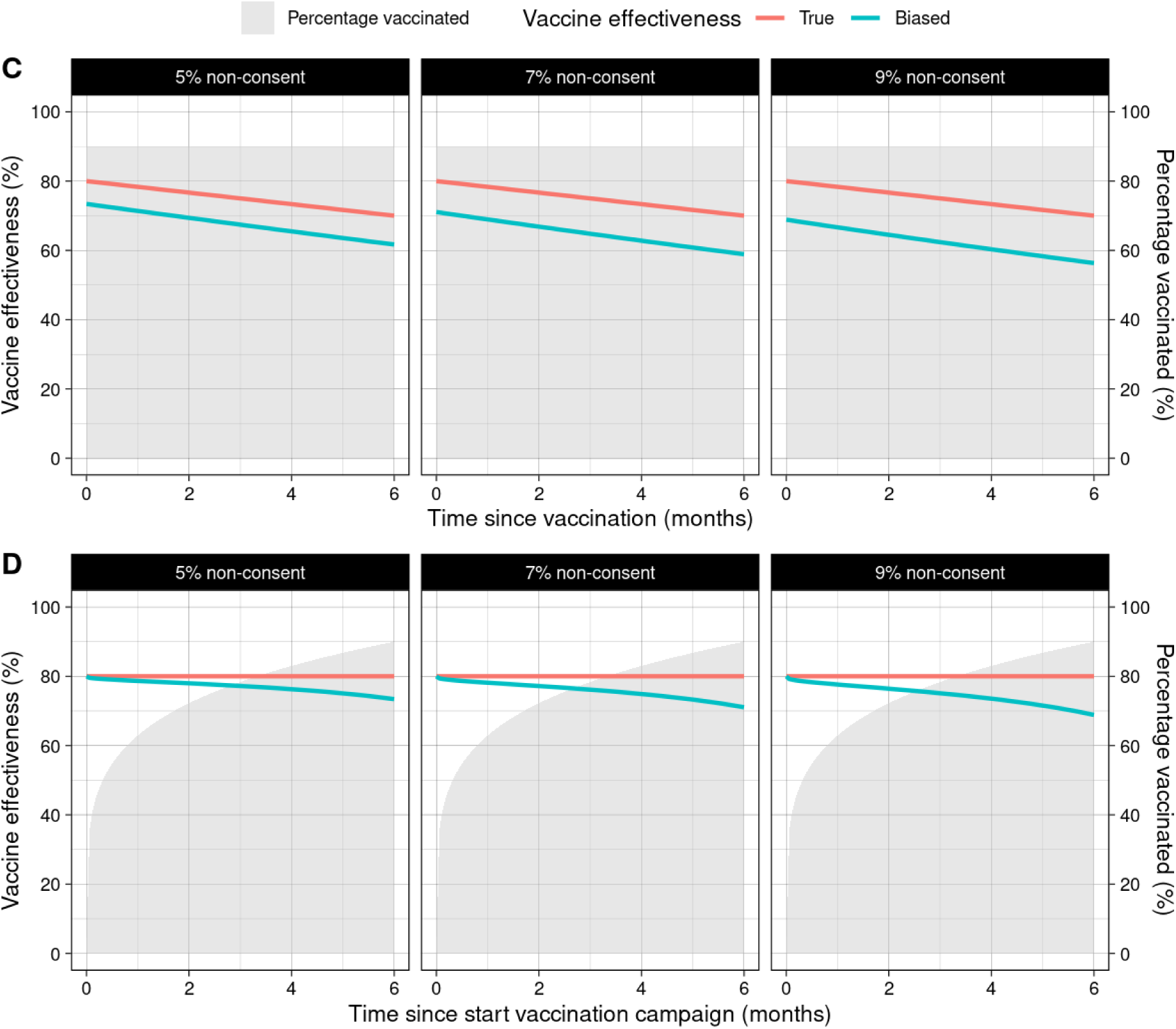
Illustration of theoretical bias by true vaccine effectiveness, vaccination uptake and non-consent percentage. A: Relative bias of the relative risk; B: Absolute bias of the vaccine effectiveness; C: Small overestimation of VE waning in case of constant vaccination coverage and true waning over time; D: Suggestion of VE waning due to increasing vaccination coverage over time when true VE is constant. VE: vaccine effectiveness. Gray areas in panel C and D indicate the true vaccination uptake in the population.

Holding the true VE and non-consent percentage constant, an increase in vaccination coverage (e.g. at the start of the vaccination campaign) led to a substantial bias, which would erroneously suggest waning of the VE (Figure 2D).

### Description of vaccination records and consent percentages in the dataset

From the total Dutch population aged 12 years and above, covariate date was missing in 0.1% of the records from the Dutch Personal Records Database, 0.7% of the CIMS records, 0.5% of the anonymous GGD dataset, and 4.1% of records in the NICE dataset. Missing records from NICE related mostly to the inability to link to the CIMS registry due to a missing or incorrect national identification number (3.9%). The analyzed population consisted of 15,504,106 individuals, of which 7,826,174 (50.5%) were female. 8,143,074 (52.5%) were aged 12-49 years, 4,747,165 (30.6%) 50-69 years, and 2613867 (16.9%) 70+ years. At the end of the follow-up period, 5,453,019 (67.0%), 3,999,842 (84.3%), and 2,270,686 (86.9%) individuals in the age groups 12-49, 50-69, and 70+ years, respectively, had a linked vaccination record in CIMS (Table 1). The number of persons who had been vaccinated but did not consent for registration in CIMS, based on the GGD data only (partially-corrected dataset), was 429,177 (7.3%), 210,338 (5.0%), and 151,909 (6.3%) for these same age groups. Regression model predictions of consent percentage by age, sex, region, and calendar time are visualized in Supplementary Figure S1 and S2. After correction using the second imputation approach (fully-corrected dataset), the non-consent percentages increased to 461,064 (7.8%), 302,377 (7.0%), and 169,882 (7.0%), respectively (Table 1). The vaccination uptake and estimated proportion misclassified over time is depicted in Figure 3. Misclassification, expressed as the proportion of persons originally classified as non-vaccinated, was highest in the 70+ age group.

**Figure 3:**
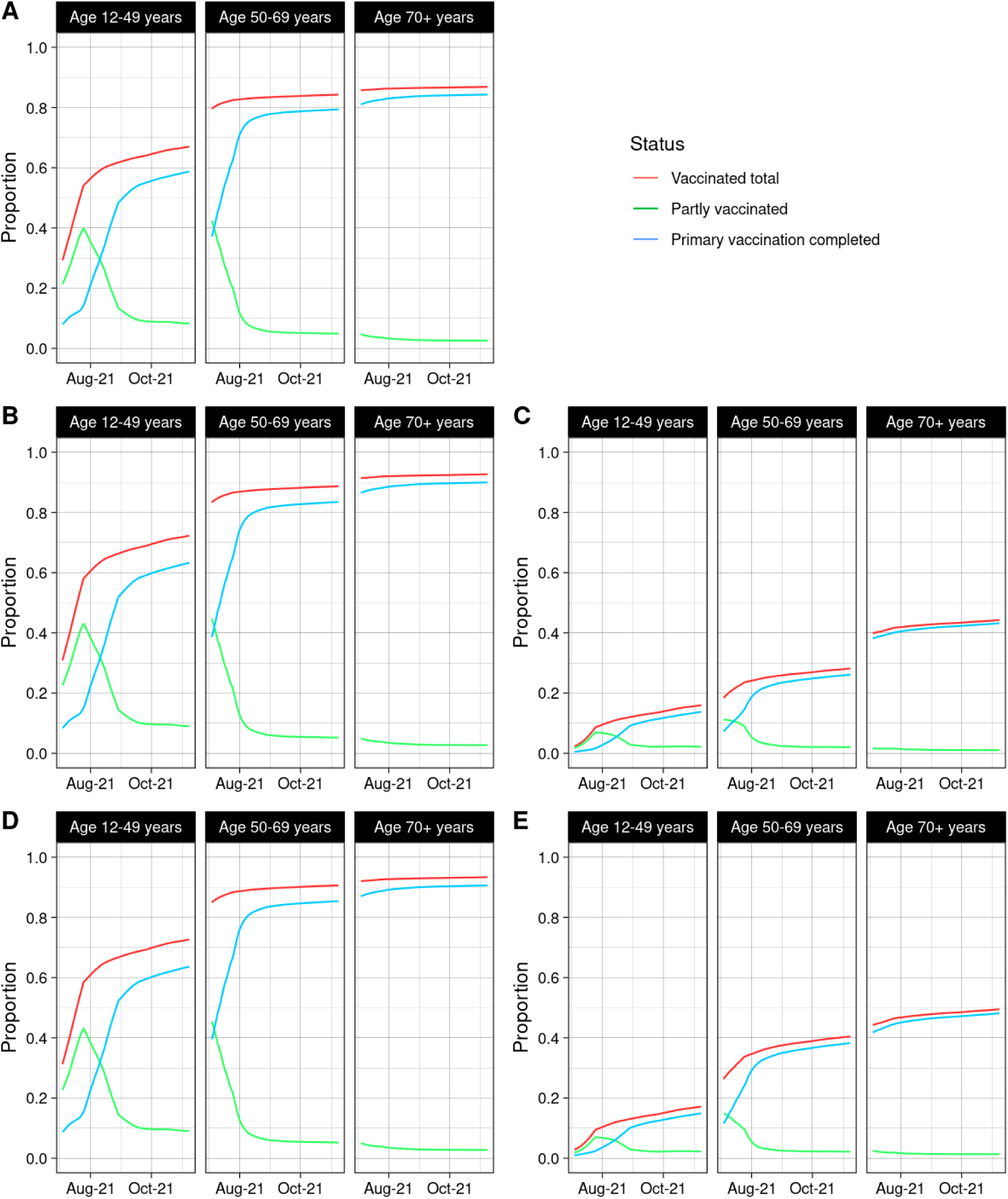
Misclassification of vaccination status over time by age group. A: Vaccination status in the original dataset; B: Vaccination status in the partially-corrected dataset; C: Vaccination status in the partially-corrected dataset among persons originally classified as unvaccinated; D: Vaccination status in the fully -corrected dataset; E: Vaccination status in the fully-corrected dataset among persons originally classified as unvaccinated.

**Table 1:**
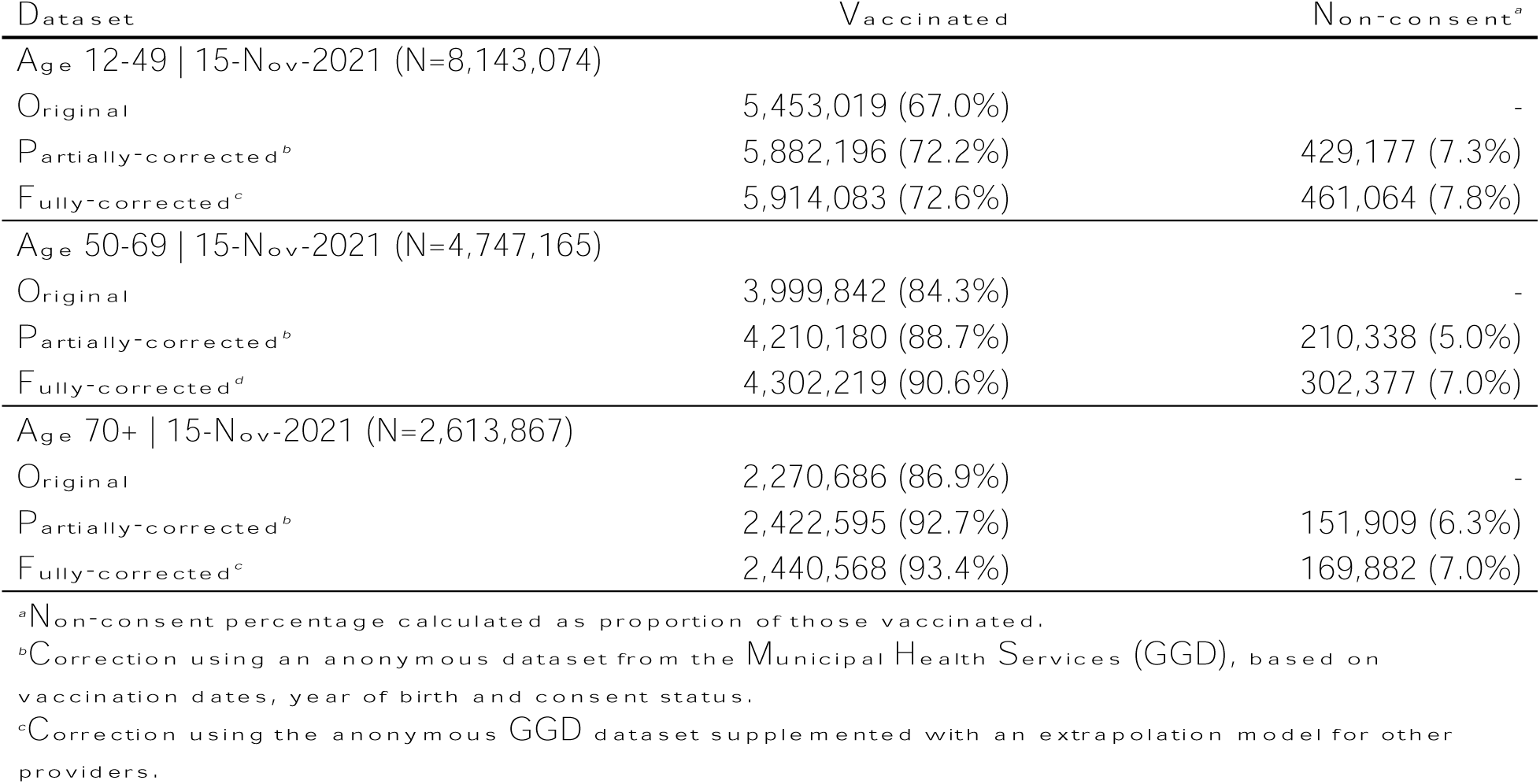
Vaccination status at the end of the study period and estimated non-consent percentage by dataset and age group.

| **Dataset** | **Vaccinated** | **Non-consent<sup>a</sup>** |
| --- | --- | --- |
| Age 12-49 \| 15-Nov-2021 (N=8,143,074) |  |  |
| Original | 5,453,019 (67.0%) | - |
| Partially-corrected<sup>b</sup> | 5,882,196 (72.2%) | 429,177 (7.3%) |
| Fully-corrected<sup>c</sup> | 5,914,083 (72.6%) | 461,064 (7.8%) |
| Age 50-69 \| 15-Nov-2021 (N=4,747,165) |  |  |
| Original | 3,999,842 (84.3%) | - |
| Partially-corrected<sup>b</sup> | 4,210,180 (88.7%) | 210,338 (5.0%) |
| Fully-corrected<sup>d</sup> | 4,302,219 (90.6%) | 302,377 (7.0%) |
| Age 70+ \| 15-Nov-2021 (N=2,613,867) |  |  |
| Original | 2,270,686 (86.9%) | - |
| Partially-corrected<sup>b</sup> | 2,422,595 (92.7%) | 151,909 (6.3%) |
| Fully-corrected<sup>c</sup> | 2,440,568 (93.4%) | 169,882 (7.0%) |

<sup>a</sup>Non-consent percentage calculated as proportion of those vaccinated.
<sup>b</sup>Correction using an anonymous dataset from the Municipal Health Services (GGD), based on vaccination dates, year of birth and consent status.
<sup>c</sup>Correction using the anonymous GGD dataset supplemented with an extrapolation model for other providers.

### Corrected VEs

All models converged after fewer than 5 iterations (Supplementary Figure S3). Both the estimated absolute bias of the VE (Figure 4, supplementary table S1) and the estimated relative bias of the RR (Supplementary Figure S4 and Table S1) were largest for the 70+ years age group. In the sensitivity analysis, assuming a lower risk of COVID-19 hospital or ICU admission for the non-consenting vaccinated individuals, relative to the consenting vaccinated individuals, yielded a larger bias in vaccine-effectiveness estimates; conversely, a higher risk of the endpoint for non-consenting individuals yielded a smaller bias (Supplementary Figure S4 and Table S1). The estimated fraction of information missing for the log(RR) (γ_mis_), i.e. the relative loss of precision of the method to estimate the corrected RR as compared to the hypothetical scenario in which there is no misclassification of vaccination status, was between 0.05 and 0.17, depending on the age group and imputation approach. This corresponds to a statistical efficiency of 95% and 83%, and an approximately 3% and 10% relative increase of the width of the confidence interval, respectively (see Supplementary Table S2).

**Figure 4:**
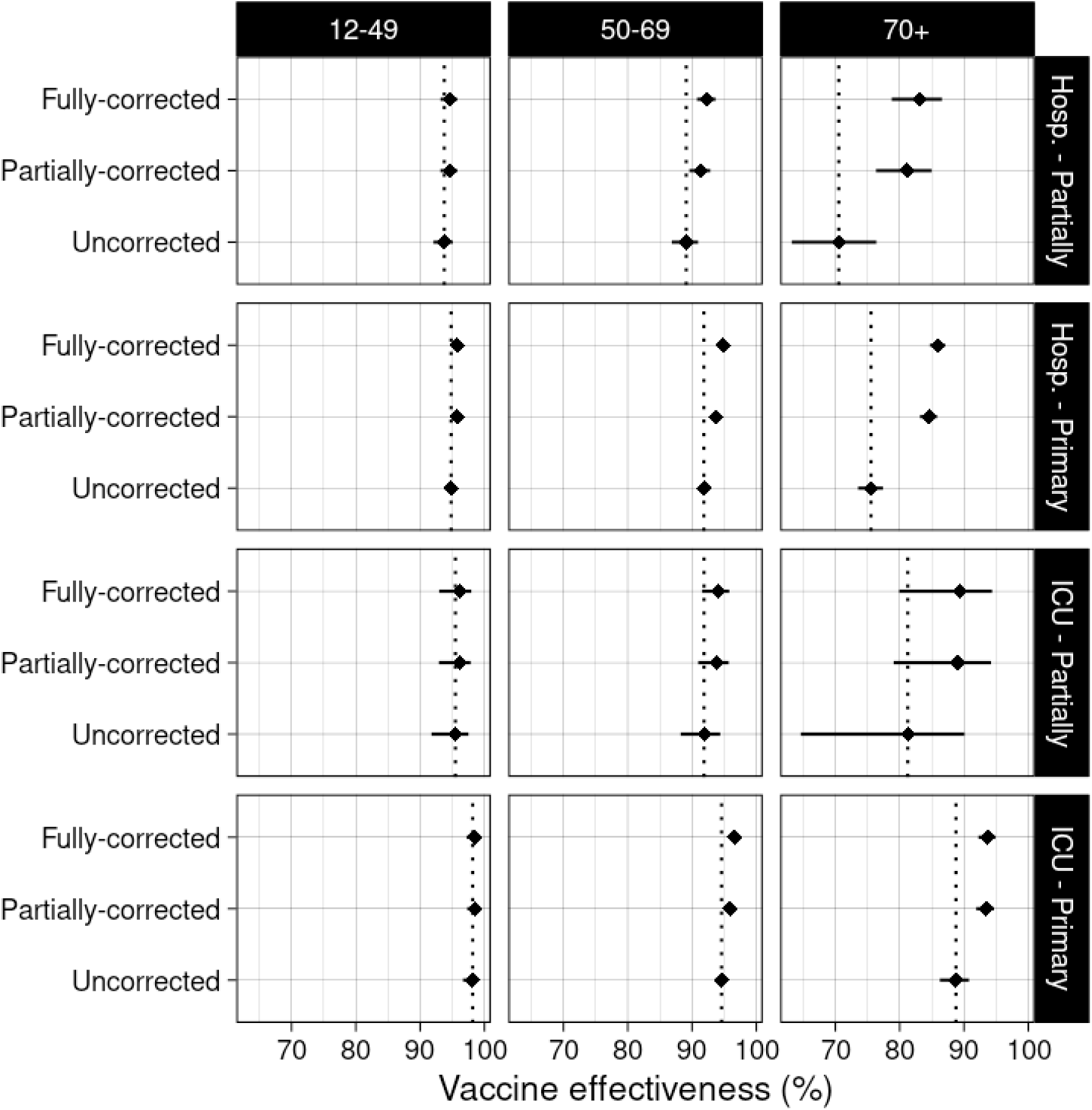
Original and corrected VE estimates assuming the risk of acquiring the endpoint is independent of providing consent. Abbreviations: hosp: hospitalization (including ICU admissions); ICU: intensive care unit admission.

## Discussion

It is known that non-differential (i.e. random) misclassification of exposure status generally causes a bias towards zero (i.e. towards no effect) although some conditions such as a polytomous exposure variable can lead to a bias away from zero.(7) In our setting, in which misclassification is always to the non-vaccinated reference group, a bias towards zero can be expected. Yet, the results of the current study caution to not ignore relatively small fractions of misclassification of exposure. For the 70+ years population, in which 7.0% of vaccinated individuals were estimated to be misclassified as non-vaccinated due to non-consent, the bias was -10 percentage points for the VE against hospitalization and -5 percentage points for the VE against ICU admission. For monitoring of vaccine effectiveness, time trends and between-group differences are highly important as they can signal (age-specific) waning immunity, or a lower VE against emerging virus variants. Accurate estimation of the VE is therefore important to support policy decisions, for example about the need of booster vaccinations.

Over the past decades different statistical approaches have been published to estimate or correct bias due to misclassification in exposure status. Probabilistic sensitivity analyses can be used to present (a range of) bias-corrected effect estimates based on (a range of) assumptions about the risk of misclassification of the exposure variable.(8) With this method, exposure status of individual records is reclassified stochastically based on the assumed parameters which may be based on data or best educated guess. Multiple imputation, regression calibration, and maximum likelihood estimation of the joint model of misclassification and outcome of interest are validated methods for the correction of misclassification of exposure; these models require that exposure is measured without error in a subset of the population.(9, 10) The misclassification simulation extrapolation (MC-SIMEX) method can be used to reduce bias due to misclassification if the misclassification matrix is known or estimated externally.(11) Bayesian correction models for misclassification have also been proposed.(12) More recent developments include the use of generalized linear finite mixture models, reparameterized imputation for measurement error, and sensitivity-specificity imputation to handle misclassification of exposure status.(13–15) These models are applicable to individual level data and utilize a pattern of exposure misclassification related to exposure and outcome status based on either a subset of the study, external data, or assumptions. They are not directly applicable to our study, because separate data streams generated aggregated numerator (admissions) and denominator (population) data, while information about non-consent was available for almost the entire denominator but not for the numerator. Therefore, we had to develop a customized model using the same assumption that all of the above methods apply: that the true determinant status can be predicted based on the observed status and patterns of misclassification, and an additional assumption that the VE was similar for consenting and non-consenting vaccinated individuals, conditional on the covariates in the model. Our method can be generalized to similar situations where information about the amount of misclassification is available for (a subset of) either the numerator or the denominator.

The corrected VE estimates that we obtained in our study are in line with VE estimates against COVID-19 hospitalization observed in other studies conducted during the Alpha and Delta dominant periods.(16) The extent to which these studies suffer from similar and/or other forms of bias is difficult to assess due to lack of reporting. The large heterogeneity in vaccine effectiveness observed between studies, giving I^2^ values >90% in most of the pooled analyses in a meta-analysis of observational studies, suggests that methodological heterogeneity is present.(16) Clinical heterogeneity may also play a role but is expected to be limited when comparing the same age group between comparable countries.

We demonstrated that as vaccination uptake increased, assuming that a relatively constant proportion of vaccinated individuals decline to give consent for registration of their vaccination, the misclassification bias also increased. As a result, time trends are more difficult to interpret. In fact, trends of decreasing VE over time may be interpreted as vaccine waning and lead to recommendations for earlier booster vaccination. In the Netherlands this was presumably not the case as vaccination uptake was relatively stable over the period in which a decreasing VE was observed. However, this may not be the same elsewhere or in the future. We also demonstrated that the magnitude of bias differed markedly between young people and older adult populations, which can lead to incorrect conclusions about age-differences in VE, especially because such differences are biologically plausible. The larger bias in the 70+ years population is mostly due to a high vaccination uptake in this age group; as a result, a larger proportion of those classified as being unvaccinated are in reality vaccinated. Clearly, it cannot simply be assumed that bias due to misclassification will be the same over time or between subgroups. Our results demonstrate the importance of complete vaccination data for monitoring of effectiveness of vaccination programs. In addition, for epidemiological studies and surveillance where this may not always be possible, they call for a discussion on how to routinely collect anonymous data of non-consenting individuals and implement misclassification correction models.

The availability of anonymous COVID-19 vaccination data by calendar date, birth year, sex and region from the GGD, which administered the vast majority of COVID vaccinations in the Netherlands, was essential to quantify the information bias resulting from the informed consent procedure. The estimated statistical efficiency of the method, compared to having 100% consent, was reasonably high. Therefore, if the model assumptions - in particular non-differential misclassification - are correct, and non-consent percentages are reasonably low, the bias can be relatively well corrected.

Recent implementation of informed consent in the Dutch childhood vaccination program resulted in a high non-consent percentage of 12%.(17) Combined with the high vaccination uptake this will result in relatively large bias in VE estimates. The use of biased VE estimates may have undesirable consequences for vaccination policy. Although the method, developed as part of this study, could be used to correct the information bias also for other vaccination programs, there are some limitations. First, the model relies on the assumption of non-differential misclassification, i.e. that vaccinated persons with and without providing consent have a similar incidence of the disease of interest conditional on their covariates. This is difficult to verify within our study and only sensitivity analyses could shed light on the importance of this assumption. In case of differential misclassification, higher non-consent percentage will result in a higher impact of misspecification of this assumption. When deemed important, the presence, direction and magnitude of differential misclassification and other important parameters of the misclassification correction model could be estimated using a survey. For example, a survey of 1209 parents in the US to compare parent-reported childhood vaccination status to EHR data found 94% agreement for receiving no vaccination and 87% for receiving all vaccinations with no delay.(18) However, a survey approach may also not be unbiased when misclassifications is due to non-consent.

Second, higher non-consent percentages such as those currently observed in the Dutch childhood vaccination program will also increase the statistical uncertainty of the model, reflected in wider confidence intervals. Finally, in the current study we were able to carry out imputations based on covariate-specific non-consent percentages that covered 86% of vaccinations. Critical information for non-consenting individuals is the (approximate) vaccination date, number of previous vaccinations, and covariates relevant to the disease under study. Unfortunately, no covariate-specific consent data are available for the Dutch childhood vaccination program. Although a global non-consent percentage could also be used, such an approach will result in less reliable imputations.

Several other limitations of the study need to be addressed. First, an anonymous dataset with informed consent status was only available for the GGD and not for other health care providers administering COVID-19 vaccines, such as general practitioners and hospitals. In the partially-corrected analysis we conservatively assumed that other providers had consent percentages of 100%, thereby underestimating the bias. The fully-corrected analysis assumed that consent percentages were similar between GGD and other providers conditional on birth year, sex, region, and calendar date. Individuals aged 50-69 years were relatively more often vaccinated by other providers. Within this age group, people born in 1956-1960 were initially invited to be vaccinated by their general practitioner. This can be appreciated from a larger difference between the corrected estimates from the fully-corrected compared to the partially-corrected analyses.

Second, the current analysis only corrects VE for misclassification of vaccination status. Other biases may be present including unmeasured confounding, as the surveillance data does not include data on comorbidities, socio-economic status, behavior or previous infections. For example, if individuals at increased risk of COVID-19 hospitalization because of comorbidity are vaccinated more often, this will result in confounding bias towards zero.(19) Determinants of healthy vaccinee bias are much more difficult to observe and may result in confounding in the opposite direction as they often related to behavioral aspects. Another form of information bias that may have occurred as part of the COVID-19 hospitalizations concern admissions for a reason not related to COVID-19 where SARS-CoV-2 infection was a secondary diagnosis. Such misclassification of the outcome most likely causes a bias towards a lower VE. To the best of our knowledge, secondary SARS-CoV-2 infection diagnoses were less of an issue during the Delta period than after the emergence of Omicron so it may not affect our analysis that much. As different biases may be in different directions, the current study serves mostly to quantify the amount of bias from one source, rather than claiming to provide an unbiased estimate of the true VE.

A third limitation is that we have not analyzed the VE by time since vaccination in our model and did not include periods where booster vaccinations were available. The method presumably is less stable when time since vaccination and other vaccination statuses are incorporated, as this would result in more uncertainty about the correct classification of hospitalized individuals with no linked vaccination records and hence in lower precision of the corrected VE estimates. The end date of this analysis coincided with the emergence of the omicron variant. The higher VE waning rates associated with this variant would warrant incorporation of time since vaccination in the analysis. Reasonable additional assumptions such as that VE can only decrease by time since vaccination may help stabilize the imputation of time since vaccination. Contemporary VE analyses may suffer less from misclassification because they often compare seasonal booster vaccines to previously vaccinated individuals. As consent for the primary series is a strong predictor of consent for the booster (personal communication RIVM), restriction of the population to those that ever consented reduces the amount of misclassification of the booster vaccine.

In conclusion, relatively small proportions of misclassified vaccination status due to the informed consent procedure implemented in the Netherlands resulted in a substantial downward bias of the VE estimates for COVID-19 hospitalization and ICU admission and potentially in incorrect conclusions about changes in the VE over time or difference in the VE between subgroups. Covariate-specific consent data should be available such that a model that takes into account misclassification can be used to correct for this form of bias, but these rely on a constant risk assumption which may not hold true. Future facilitation of the use of routinely collected health data, while protecting privacy rights and personal autonomy, is crucial to increase its value in surveillance and research for public health.

## Data Availability

All data produced in the present study are available upon reasonable request to the authors

## Statements and Declarations

### Author Contributions

Conceptualization: CHvW, MJK, BdG. Data analysis: CHvW. Data analysis verification: SM. Supervision: MJK. Writing original draft manuscript: CHvW. Critical review of the manuscript: all authors.

### Ethics approval

This is a secondary analysis of another study (https://doi.org/10.1101/2022.07.21.22277831) for which the research proposal was assessed by the Centre for Clinical Expertise at the RIVM. They verified whether the work complies with the specific conditions as stated in the law for medical research involving human subjects (WMO), and were of the opinion that the research does not fulfill one or both of these conditions and therefore conclude it is exempted for further approval by the ethical research committee.

### Consent to participate

Not applicable to this type of research.

## References

1. Hahné S, Bollaerts K, Farrington P. Vaccination Programmes: Epidemiology, Monitoring, Evaluation: Taylor & Francis; 2021.

2. Archive COVID-19 vaccination figures 2022. RIVM, Bilthoven. https://www.rivm.nl/en/covid-19-vaccination/archive-covid-19-vaccination-figures-2022. Accessed 12-Jul-2022.

3. [Vaccine effectiveness in preventing hospital and ICU admission in the Netherlands]. RIVM, Bilthoven. https://www.rivm.nl/sites/default/files/2021-11/Analyse%20VE_Update%20-%20versie%2018%20nov%202021-FINAL-2.docx.

4. Gier Bd, Kooijman M, Kemmeren J, et al. COVID-19 vaccine effectiveness against hospitalizations and ICU admissions in the Netherlands, April-August 2021. medRxiv. 2021:2021.09.15.21263613. doi:10.1101/2021.09.15.21263613

5. Team RC. R: A language and environment for statistical computing. Vienna, Austria: R Foundation for Statistical Computing; 2021.

6. Wood SN. Fast Stable Restricted Maximum Likelihood and Marginal Likelihood Estimation of Semiparametric Generalized Linear Models. Journal of the Royal Statistical Society Series B: Statistical Methodology. 2010;73(1):3–36. doi:10.1111/j.1467-9868.2010.00749.x

7. Jurek AM, Greenland S, Maldonado G, Church TR. Proper interpretation of non-differential misclassification effects: expectations vs observations. Int J Epidemiol. 2005;34(3):680–7. doi:10.1093/ije/dyi060

8. Fox MP, Lash TL, Greenland S. A method to automate probabilistic sensitivity analyses of misclassified binary variables. Int J Epidemiol. 2005;34(6):1370–6. doi:10.1093/ije/dyi184

9. Cole SR, Chu H, Greenland S. Multiple-imputation for measurement-error correction. Int J Epidemiol. 2006;35(4):1074–81. doi:10.1093/ije/dyl097

10. Messer K, Natarajan L. Maximum likelihood, multiple imputation and regression calibration for measurement error adjustment. Stat Med. 2008;27(30):6332–50. doi:10.1002/sim.3458

11. Küchenhoff H, Mwalili SM, Lesaffre E. A general method for dealing with misclassification in regression: the misclassification SIMEX. Biometrics. 2006;62(1):85–96. doi:10.1111/j.1541-0420.2005.00396.x

12. MacLehose RF, Olshan AF, Herring AH, Honein MA, Shaw GM, Romitti PA. Bayesian methods for correcting misclassification: an example from birth defects epidemiology. Epidemiology. 2009;20(1):27–35. doi:10.1097/EDE.0b013e31818ab3b0

13. Hubbard RA, Johnson E, Chubak J, Wernli KJ, Kamineni A, Bogart A, Rutter CM. Accounting for misclassification in electronic health records-derived exposures using generalized linear finite mixture models. Health Serv Outcomes Res Methodol. 2017;17(2):101–12. doi:10.1007/s10742-016-0149-5

14. Corbin M, Haslett S, Pearce N, Maule M, Greenland S. A comparison of sensitivity-specificity imputation, direct imputation and fully Bayesian analysis to adjust for exposure misclassification when validation data are unavailable. Int J Epidemiol. 2017;46(3):1063–72. doi:10.1093/ije/dyx027

15. Edwards JK, Cole SR, Fox MP. Flexibly Accounting for Exposure Misclassification With External Validation Data. Am J Epidemiol. 2020;189(8):850–60. doi:10.1093/aje/kwaa011

16. Rahmani K, Shavaleh R, Forouhi M, et al. The effectiveness of COVID-19 vaccines in reducing the incidence, hospitalization, and mortality from COVID-19: A systematic review and meta-analysis. Front Public Health. 2022;10:873596. doi:10.3389/fpubh.2022.873596

17. Van Lier EAO, P.J.; Giesbers, H.; Hament, J-M; Van Vliet, J.A.; Drijfhout, I.H.; Hirschberg, H.; De Melker, H.E. Vaccinatiegraad en jaarverslag Rijksvaccinatieprogramma Nederland 2021: RIVM2022.

18. Daley MF, Shoup JA, Newcomer SR, et al. Assessing Potential Confounding and Misclassification Bias When Studying the Safety of the Childhood Immunization Schedule. Acad Pediatr. 2018;18(7):754–62. doi:10.1016/j.acap.2018.03.007

19. de Gier BvA, L.; Boere, T.; van Roon, A.; van Roekel, C.; Pijpers, J.; van Werkhoven, H.; van den Ende, C.; Hahné, S.; de Melker, H.; Knol, M.; van den Hof, S. COVID-19 vaccine effectiveness against mortality and risk of death from other causes after COVID-19 vaccination, the Netherlands, January 2021-January 2022. medRxiv. 2023. 10.1101/2022.07.21.22277831

