## Supplementary Appendix for "Information bias of vaccine effectiveness estimation due to informed consent for national registration of COVID-19 vaccination"

**To the manuscript titled:**

**Information bias of vaccine effectiveness estimation due to informed consent for national registration of COVID-19 vaccination: estimation and correction using a data augmentation model**

#### Index:

### Supplementary methods:

#### Derivation of bias estimation formula:

The relative risk ($RR$) is:

$$RR = 1 - VE/100$$

Taking a true incidence in unvaccinated $i_u$ yields a true incidence in the vaccinated of

$$i_v = i_u * RR$$

Assuming the incidence is independent of providing consent, we can take $i_v$ as the observed incidence in vaccinated $i'_v$ (i.e. in those providing consent) and calculate the observed incidence in unvaccinated $i'_u$ (i.e. in those not vaccinated and those not providing consent) as a weighted mean incidence of unvaccinated and non-consenting individuals:

$$i'_u = \frac{i_u * (1 - v) + i_v * v * nc}{1 - v + v * nc} = \frac{i_u * (1 - v) + i_u * RR * v * nc}{1 - v + v * nc}$$

where $v$ is the proportion vaccinated in the population and $nc$ is the proportion of vaccinated persons not providing informed consent. The observed $RR'$ is a ratio of $i'_v$ and $i'_u$. as follows:

$$RR' = \frac{i'_v}{i'_u} = \frac{i_u * RR}{\left(\frac{i_u * (1 - v) + i_u * RR * v * nc}{1 - v + v * nc}\right)} = \frac{i_u * RR * (1 - v + v * nc)}{i_u * (1 - v) + i_u * RR * v * nc}$$

The incidence parameter can be cancelled out and the formula can be simplified to:

$$RR' = \frac{RR * (1 - v + v * nc)}{1 - v + RR * v * nc}$$

#### Data sources

The NICE dataset was linked to the CIMS database using the unique national identification number to determine the COVID-19 vaccination status of hospitalized patients. Hospitalization records that could not be linked because of missing or incorrect identification numbers, as well as those with missing birth year or persons born after 2010 (i.e. >= 12 years of age in 2021), were excluded. Disease onset date was not recorded in the NICE data. To avoid attributing admissions occurring directly after vaccination to failure of the vaccine, we assumed a 7-day interval between disease onset and hospital admission, based on the median lag time in notified COVID-19 hospitalizations early in the pandemic.(1, 2) This dataset was aggregated to the daily number of hospital admissions and ICU admissions per vaccination status, birth year, sex and region (defined by the 25 national safety regions), referred to as the 'original events dataset'. For this study we selected hospitalizations between 11 July and 15 November 2021 for three reasons. First, during this period a single SARS-CoV-2 variant (Delta) was dominant. Second, VE estimates were relatively stable over this period, in contrast with later periods in which the emergence of Omicron variants was associated with a rapid decline of the VE over time.(3) Such changes in the true VE would have severely complicated our analysis. Third, booster vaccinations were administered as of 18 November 2021. Because informed consent was obtained separately for the booster vaccination, discordant consents for the primary series and the booster vaccination in CIMS would have further complicated the analysis.

The total population size on January 1st, 2021 per year of birth, sex, and region for individuals born before 2010 was obtained from the Dutch Personal Records Database, and then enriched with CIMS data to provide the daily population size per vaccination status, birth year, sex and region. This is referred to as the 'original population dataset'. Since both the original events dataset and the original population dataset use CIMS to define vaccination status, both suffer from misclassification of vaccination status (i.e. a proportion of individuals marked as unvaccinated had in reality been vaccinated).

Informed consent percentages by date, year of birth, sex, and region were based on an anonymous minimal nationwide COVID-19 vaccine register that consists of vaccination records from all individuals vaccinated by Municipal Health Services (GGDs). This register indicates whether or not informed consent for registration in CIMS was given. Based on this GGD dataset we constructed an aggregated population dataset with similar format as the original population dataset, but representing numbers of individuals who did not provide consent for registration of their primary series vaccination.

#### Statistical methods

We implemented two different imputation approaches. In the first approach we took the original population dataset as a starting point. Within each stratum of calendar time, birth year, sex and region, we substituted the number of unvaccinated persons by the same number of individuals not providing consent at the GGD with their corresponding vaccination status (partially or fully vaccinated with the primary series). This dataset is referred to as the "partially-corrected population dataset". The implicit assumption for this imputation approach is that individuals vaccinated by providers other than the GGD all provided informed consent.

In the second imputation approach, we fitted a logistic regression model for the proportion providing informed consent among individuals vaccinated by the GGD as a function of calendar time, birth year, sex, and region. The model included an interaction spline for calendar time and birth year, thus assuming smooth changes over time and age. This model was used to predict the number of non-consenting individuals among individuals vaccinated by providers other than the GGD (e.g. general practitioners or hospitals). Thus, we assumed that conditional on calendar time, birth year, sex, and region, the consent percentages were similar for GGD and other vaccination providers. We then replaced individuals classified as unvaccinated in the original population dataset with partially or fully vaccinated individuals according to the number of non-consenting individuals, stratified by calendar time, birth year, sex, and region. This dataset is referred to as the "fully-corrected population dataset". Since this is based on model prediction rather than observation, to incorporate the statistical uncertainty of this imputation, the fully-corrected population dataset was stochastically generated during each iteration (see next sections).

To illustrate the estimated amount of misclassification in the population and the change in vaccination status after correction, the prevalence of different vaccination statuses over calendar time were visually presented by age stratum for the original population dataset, the partially-corrected population dataset, and the fully-corrected population dataset. We also plotted the newly assigned vaccination statuses for individuals that were classified as unvaccinated in the original population dataset.

To model the VE, we used a Generalized Additive Model with a negative binomial link function, using number of hospital or ICU admissions as the outcome of interest and the natural logarithm of the population size as offset. Covariates included calendar time with a smooth function using a penalized thin plate regression spline with 39 degrees of freedom (k=40) and age in 5-year categories. The model yields relative risks (RR) from which the VE can be calculated as (1 – RR) * 100%; and the 95% confidence interval as (1 – upper/lower bound RR) * 100%. First, this model was fit to the original population and events dataset, thus yielding VE estimates affected by information bias due to misclassification of vaccination status.

Next, we fitted the model to the corrected events and population dataset, separately for the partially-corrected and fully-corrected datasets. However, the amount of misclassification in the original events dataset depends both on the distribution of true vaccination status and non-consent percentage in the population, and on the true VE. Therefore, we used a customized data augmentation model to impute the vaccination status stochastically. First, we determined the expected distribution of true vaccination statuses in those classified as 'unvaccinated' in the original events dataset, using the same stratification factors as for the population dataset: calendar time, birth year, sex, and region. This expected distribution was determined by multiplying the distribution of vaccination statuses in the corrected population dataset, for those classified as unvaccinated in the original population dataset, by the estimated relative risk for each vaccination status (using 1 for status 'unvaccinated'), and subsequently normalizing this to obtain a probability (see example in Figure 1). Next, we stochastically imputed the vaccination status for each hospitalized patient that was classified as 'unvaccinated' in the original events dataset. By refitting the regression model to this augmented dataset we obtained a corrected RR. Since the imputation model step uses an initial RR estimate that is biased, the newly obtained RR is not optimally corrected. Therefore, the stochastic assignment of vaccination status was repeated for a total of 10 iterations, each time using the previous RR estimate. We performed 20 independent imputations to visually inspect whether convergence was reached. This analysis was performed separately for the age groups 12-49, 50-69, and 70+ years old, the two outcome events COVID-19 hospitalization and COVID-19 ICU admission, and the two imputation methods (partially-corrected and fully-corrected).

The implicit assumption underlying the stochastic assignment of vaccination status is that the incidence of an outcome, conditional on calendar time, birth year, sex and region, is independent of providing informed consent. To assess the impact of this assumption, we performed sensitivity analyses in which we assumed that those not providing informed consent would have a higher (RR 1.5) or lower (RR 0.7) incidence compared to consenting vaccinated individuals, conditional on birth year, calendar time, sex, and region.

We calculated 95% confidence intervals for the corrected VE estimates by pooling the estimates and standard errors of the final iteration from the 20 imputations according to Rubin's rules.(4) The estimated bias was expressed as the absolute difference in percentage points of the VE and the relative difference of the RR of the bias-corrected estimates compared to the uncorrected estimates. Finally, we calculated for each scenario the relative efficiency of the data augmentation method as compared to the hypothetical scenario in which there is no misclassification of vaccination status. In multiple imputation, the fraction of observed information can be interpreted as the efficiency of the imputation model in comparison to having observed all data. Its complement, the *fraction of missing information*, can be calculated as:

$$\gamma_{mis} = \frac{B_{MI}(1 + 1/M)}{V_{MI}}$$

where $B_{MI}$ is the between-imputation variance of the log(RR) estimates, $M$ is the number of imputations, and $V_{MI}$ is the pooled variance of the log(RR). This quantity can also be used to determine the number of imputations required to obtain a stable pooled estimate.(5)

### Supplementary results

#### Figure S1: Model estimation of consent percentage for partial vaccination

By year of birth, sex, region, and calendar time (year 2021). A. B. model prediction using interaction splines for calendar time and year of birth. B. Adjusted odds ratio's for sex and region.

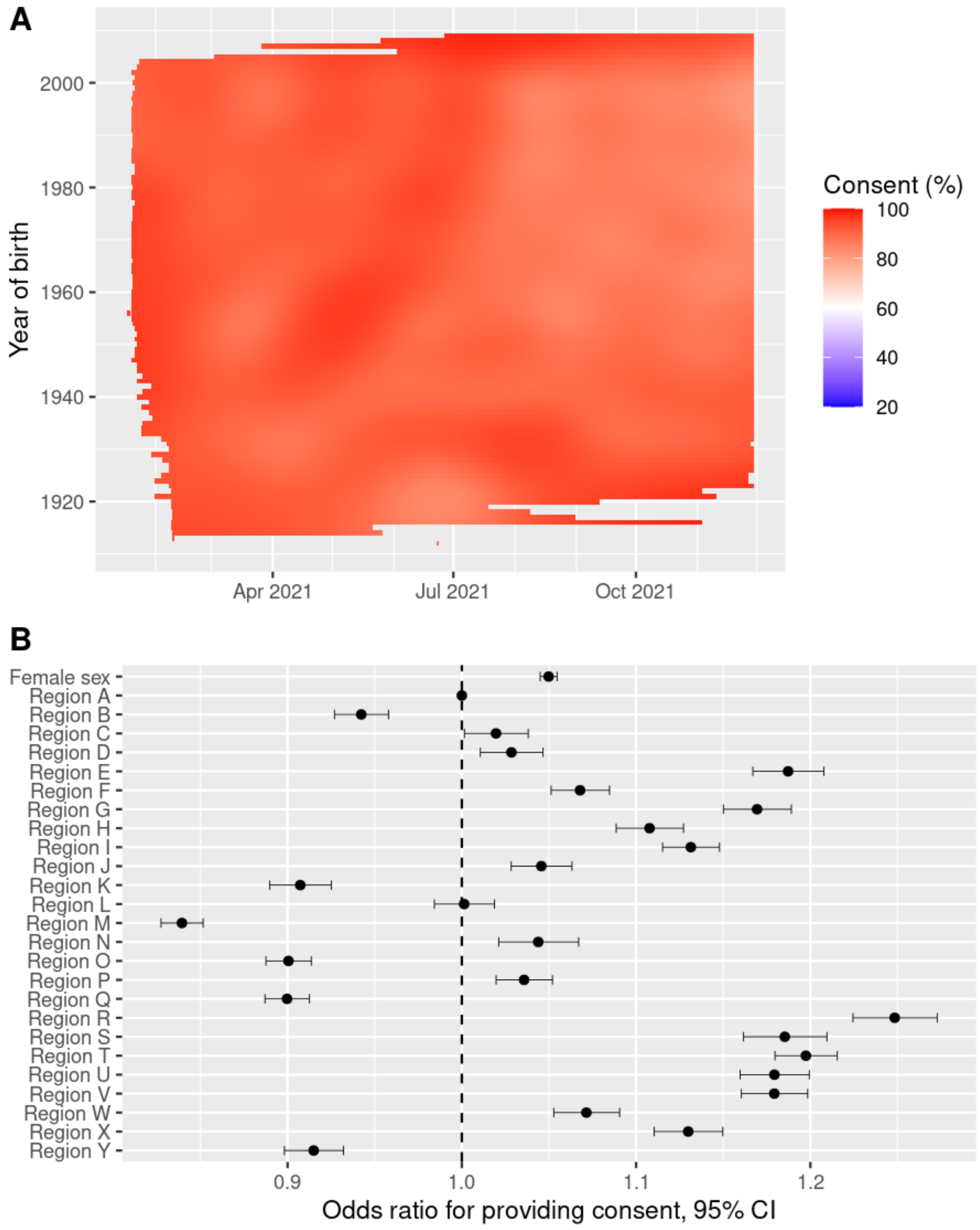

#### Figure S2: Model prediction of consent rate for full primary vaccination

By year of birth and calendar time (year 2021). A. crude rate aggregated by month and 10-year of birth. B. model prediction using interaction splines for calendar time and year of birth.

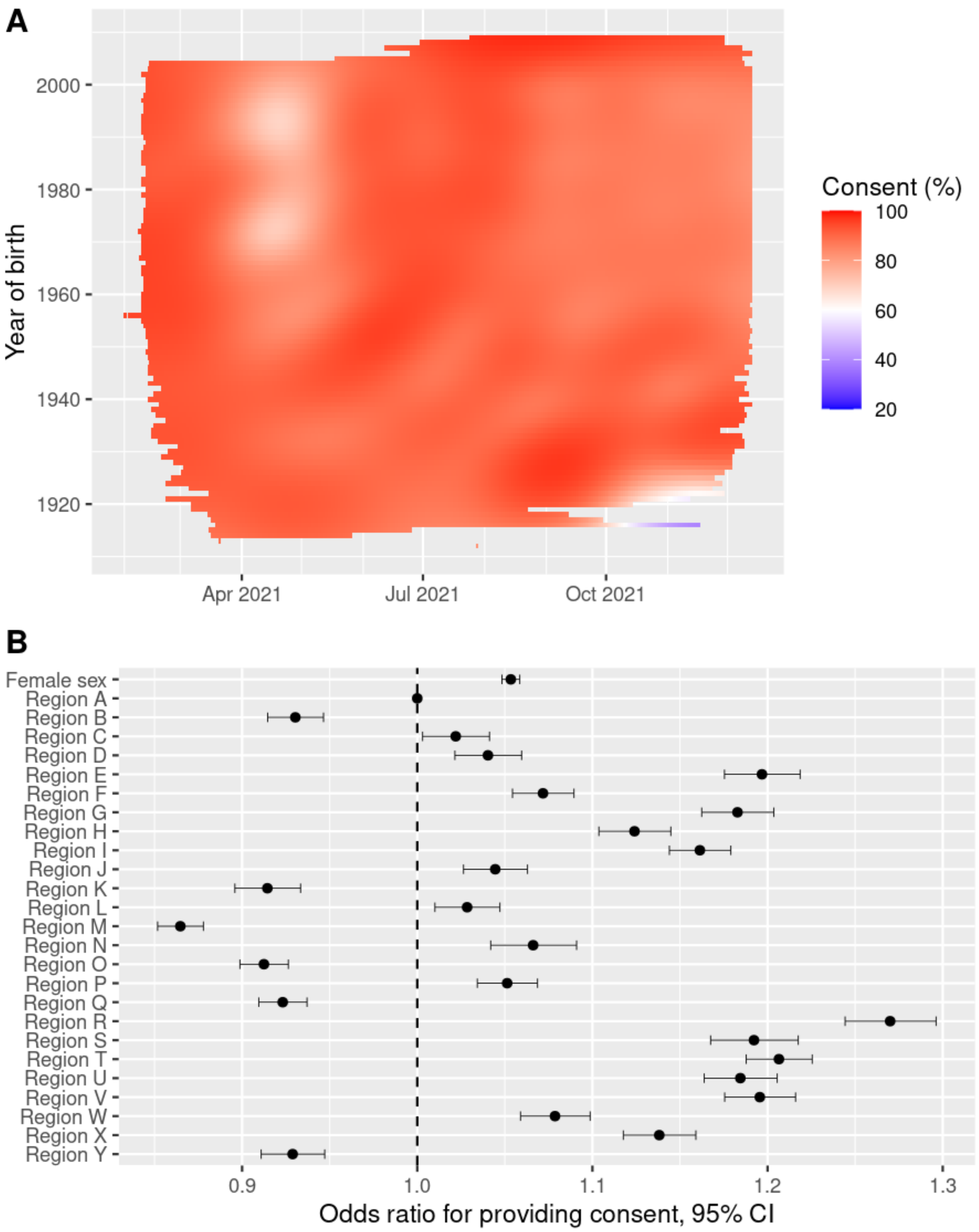

Figure S3: Convergence of models

Abbreviations: RR: relative risk, ICU: intensive care unit, GGD: municipal health service.

**MODEL 1**

Endpoint: Hospitalization, Correction: partially, RR no consent: 0.7**

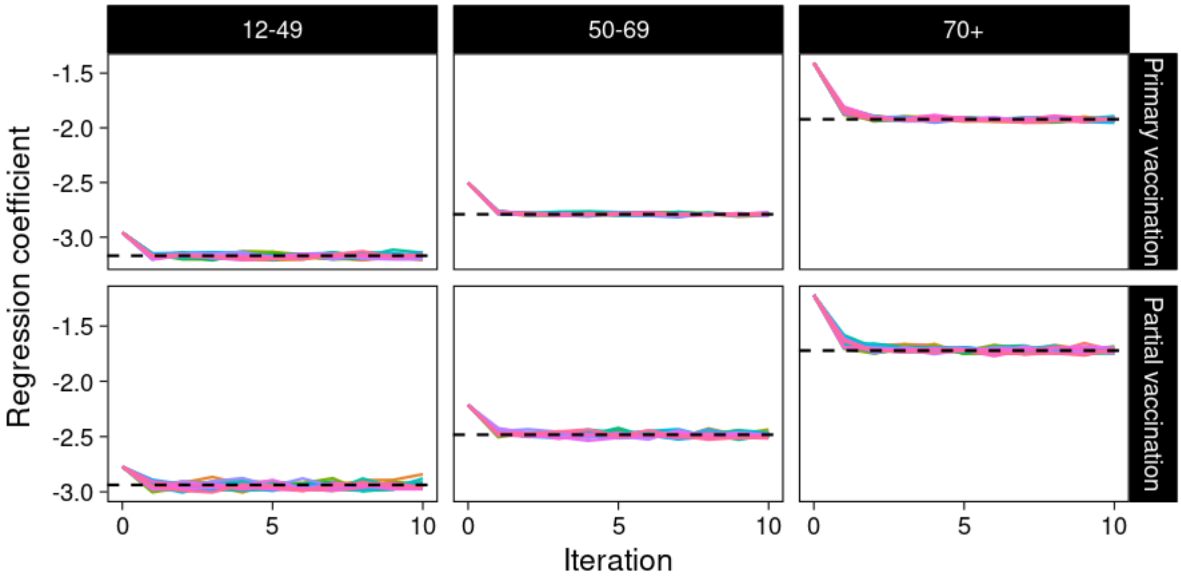

**MODEL 2**

Endpoint: Hospitalization, Fully corrected model, RR no consent: 0.7**

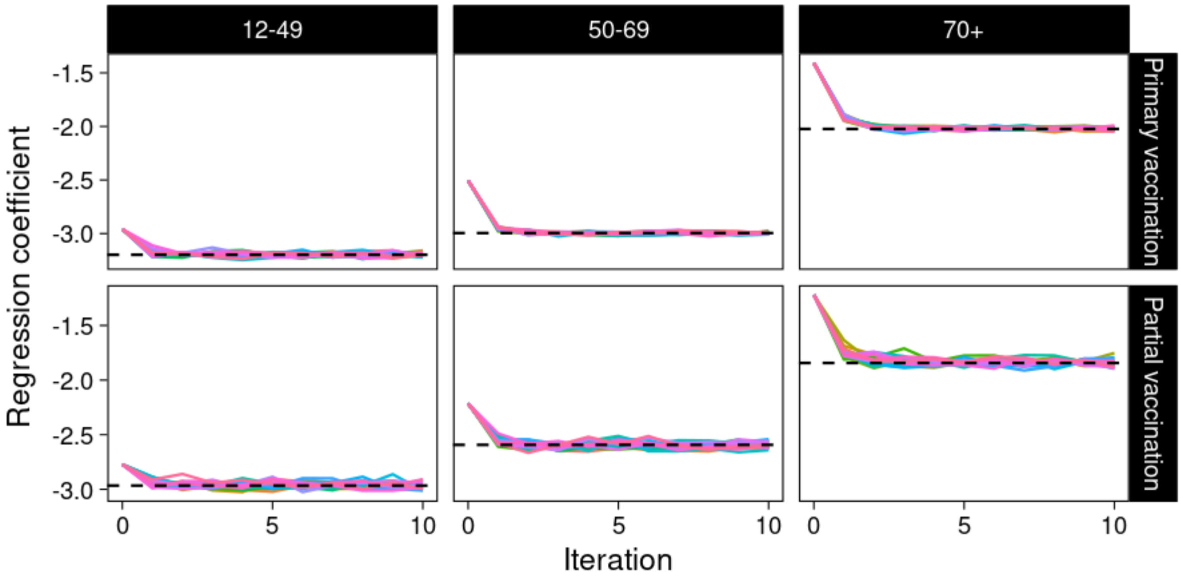

**MODEL 3**

Endpoint: Hospitalization, Partially corrected model, RR no consent: 1.0**

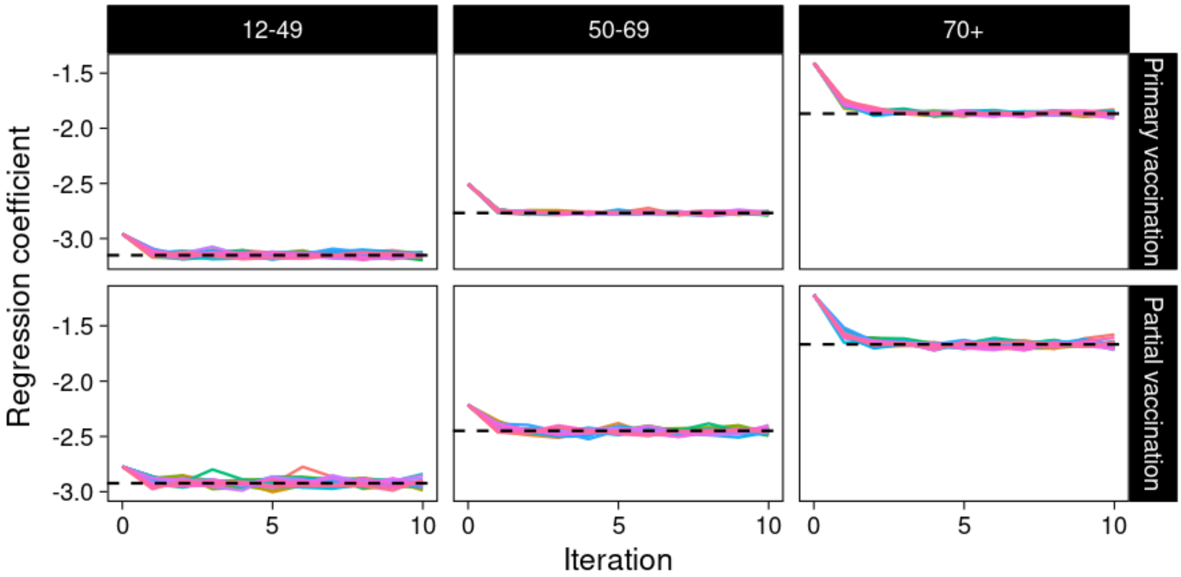

**MODEL 4**

Endpoint: Hospitalization, Fully corrected model, RR no consent: 1.0**

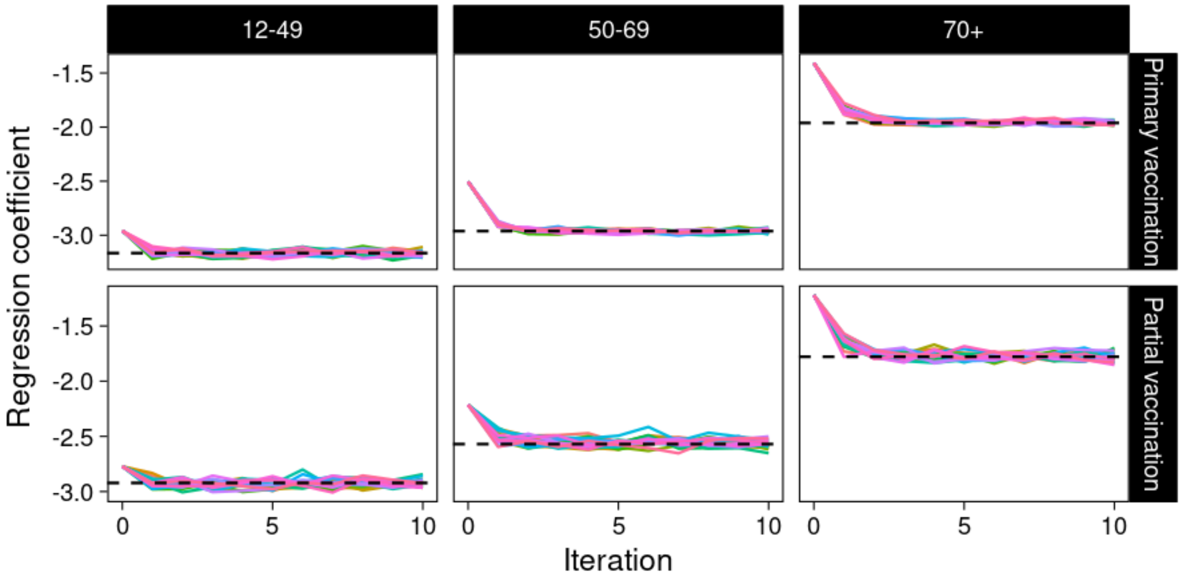

**MODEL 5**

Endpoint: Hospitalization, Partially corrected model, RR no consent: 1.5**

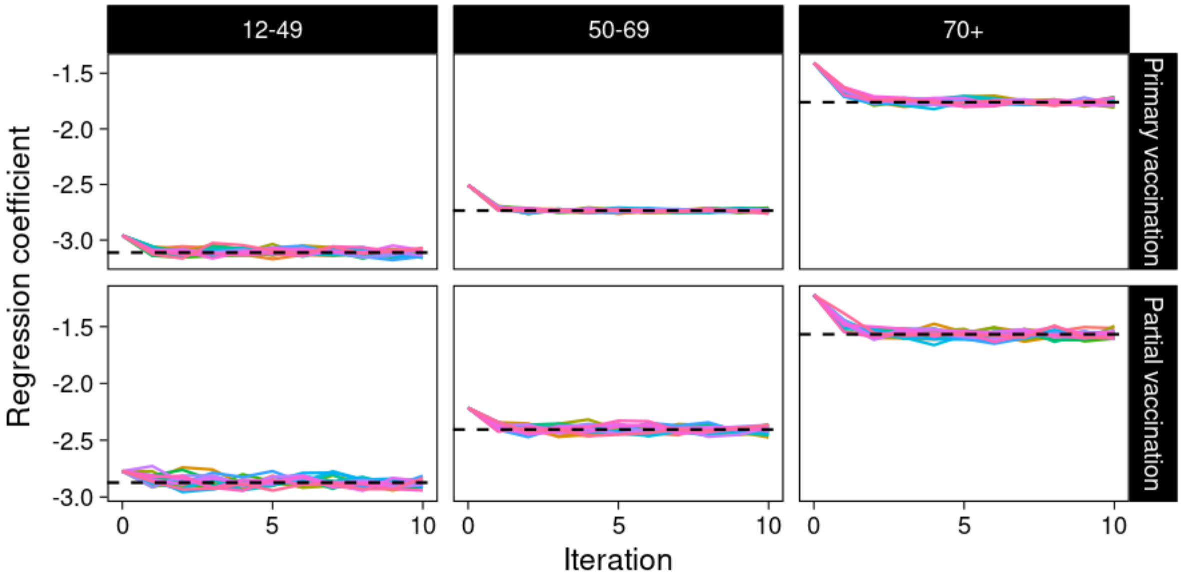

**MODEL 6**

Endpoint: Hospitalization, Fully corrected model, RR no consent: 1.5**

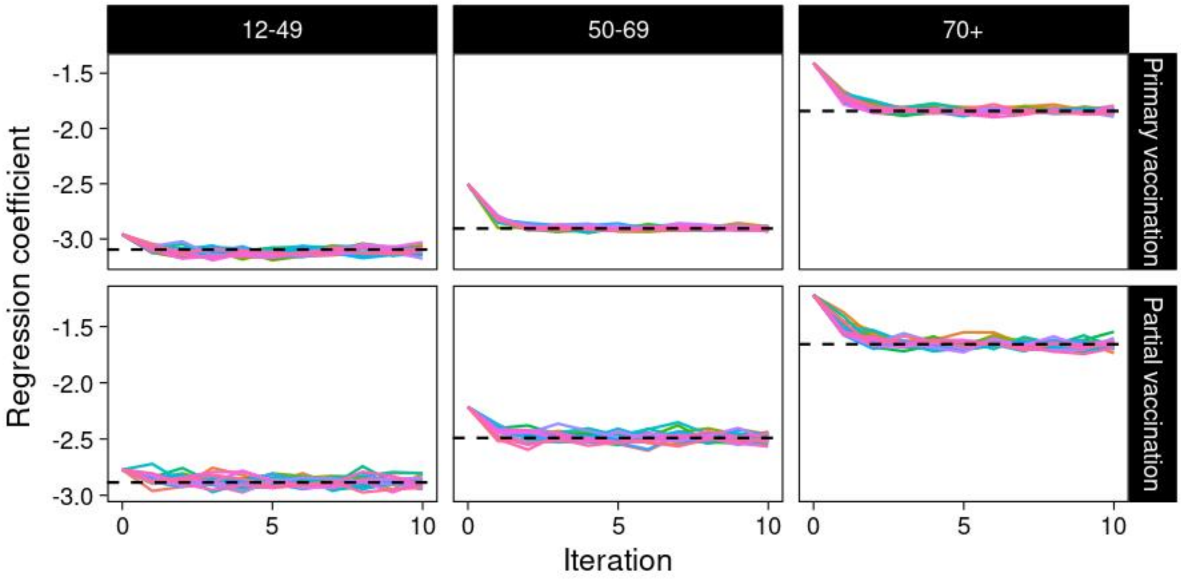

**MODEL 7**

Endpoint: ICU admission, Partially corrected model, RR no consent: 0.7**

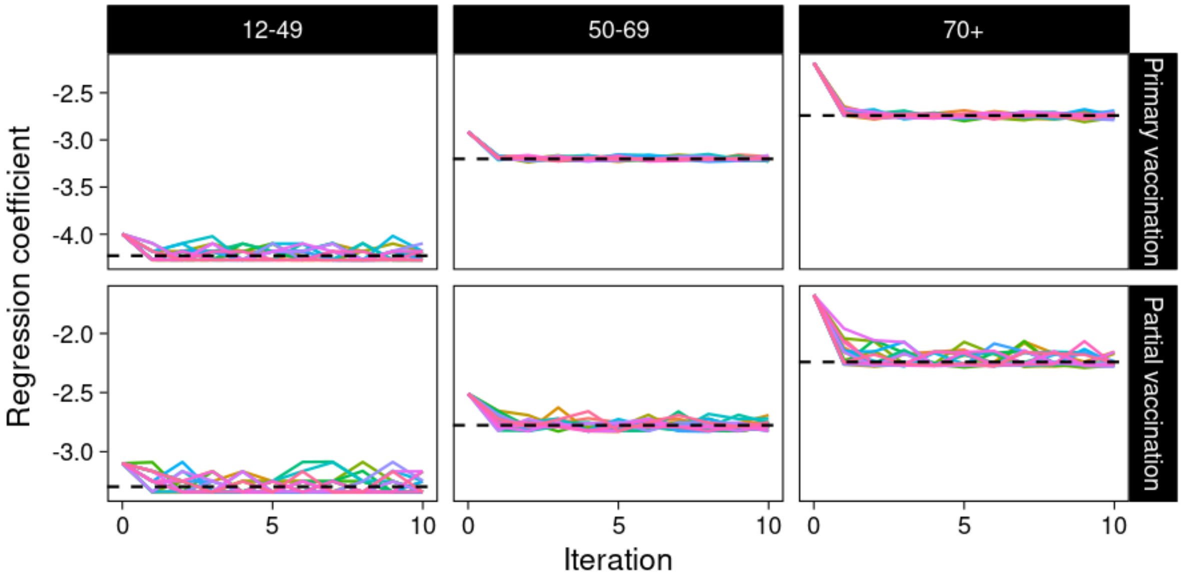

**MODEL 8**

Endpoint: ICU admission, Fully corrected model, RR no consent: 0.7**

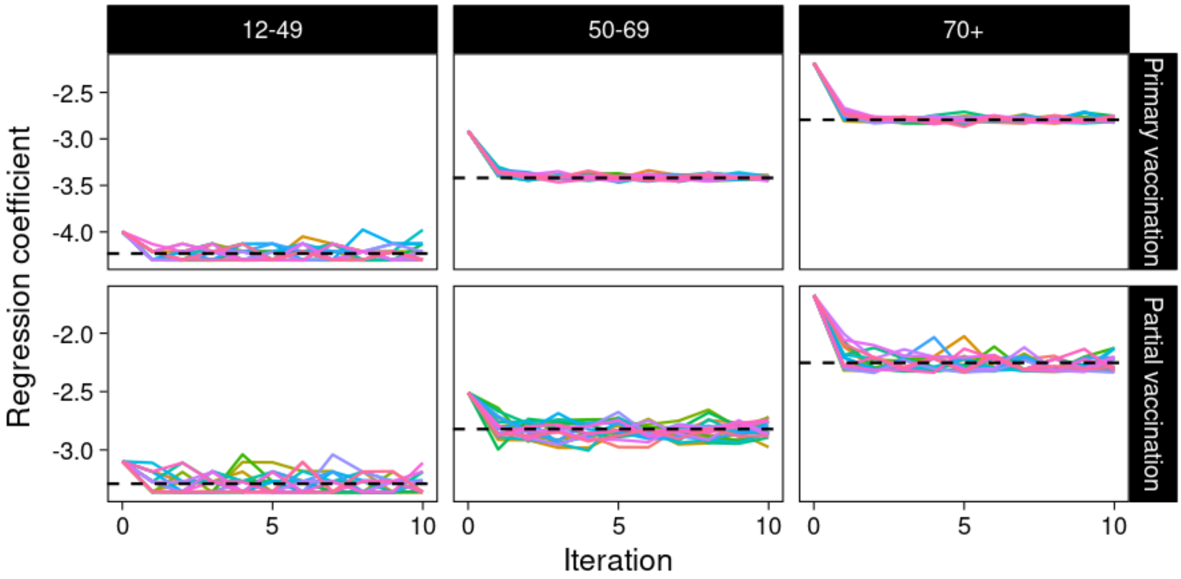

**MODEL 9**

Endpoint: ICU admission, Partially corrected model, RR no consent: 1.0**

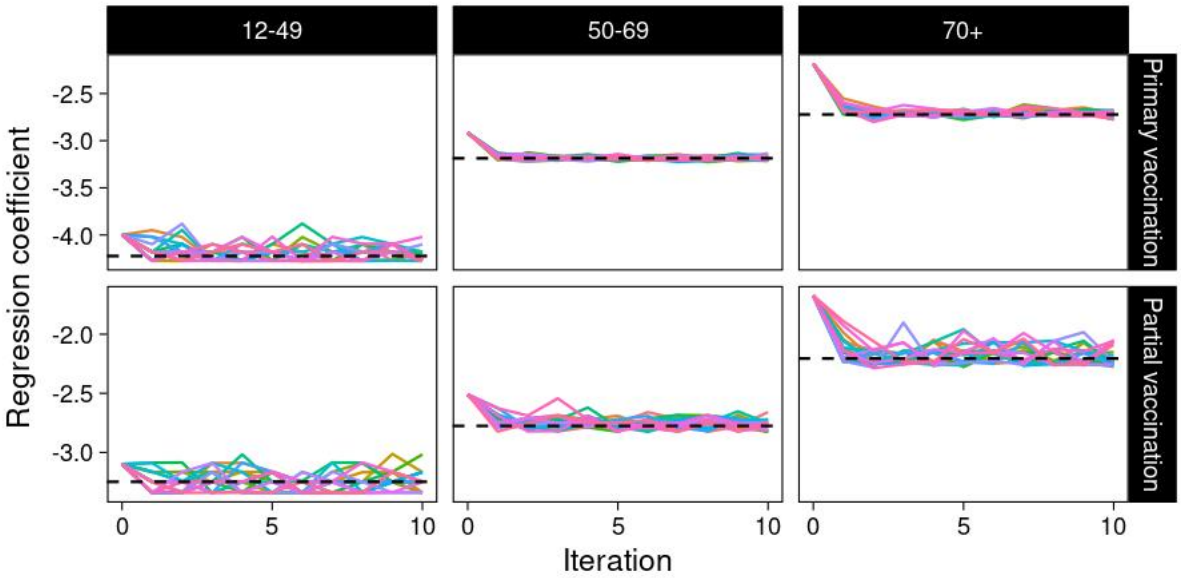

**MODEL 10**

Endpoint: ICU admission, Fully corrected model, RR no consent: 1.0**

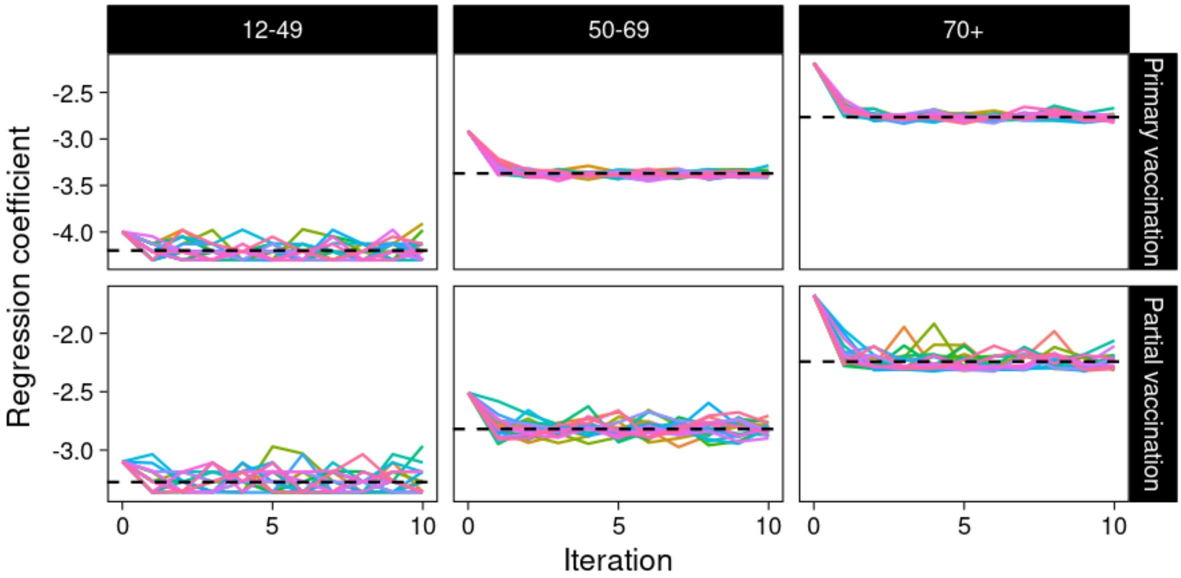

**MODEL 11**

Endpoint: ICU admission, Partially corrected model, RR no consent: 1.5**

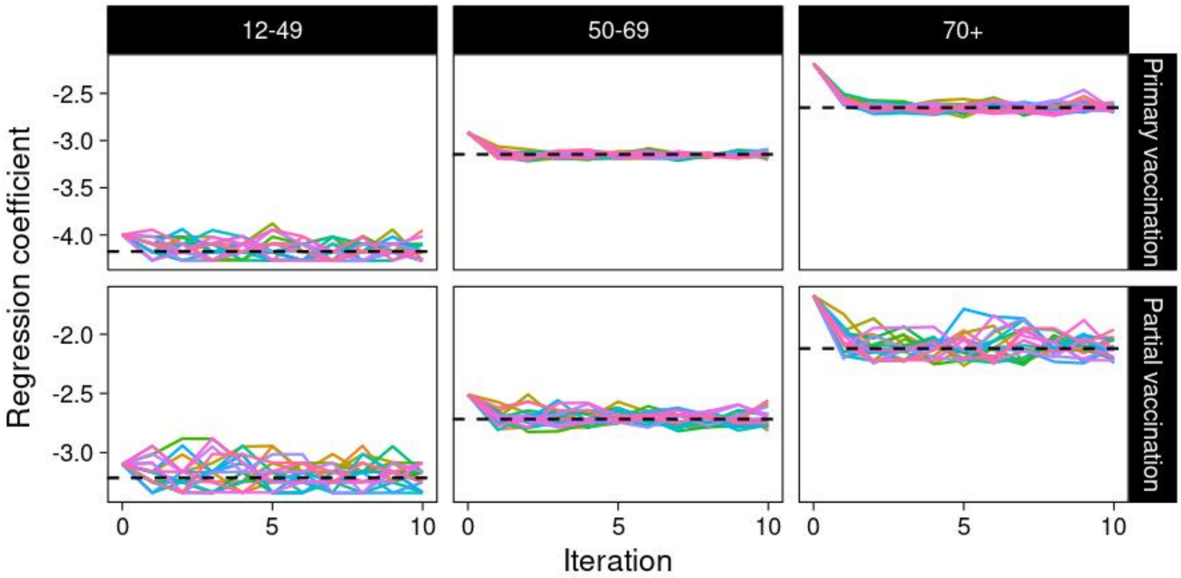

**MODEL 12**

Endpoint: ICU admission, Fully corrected model, RR no consent: 1.5**

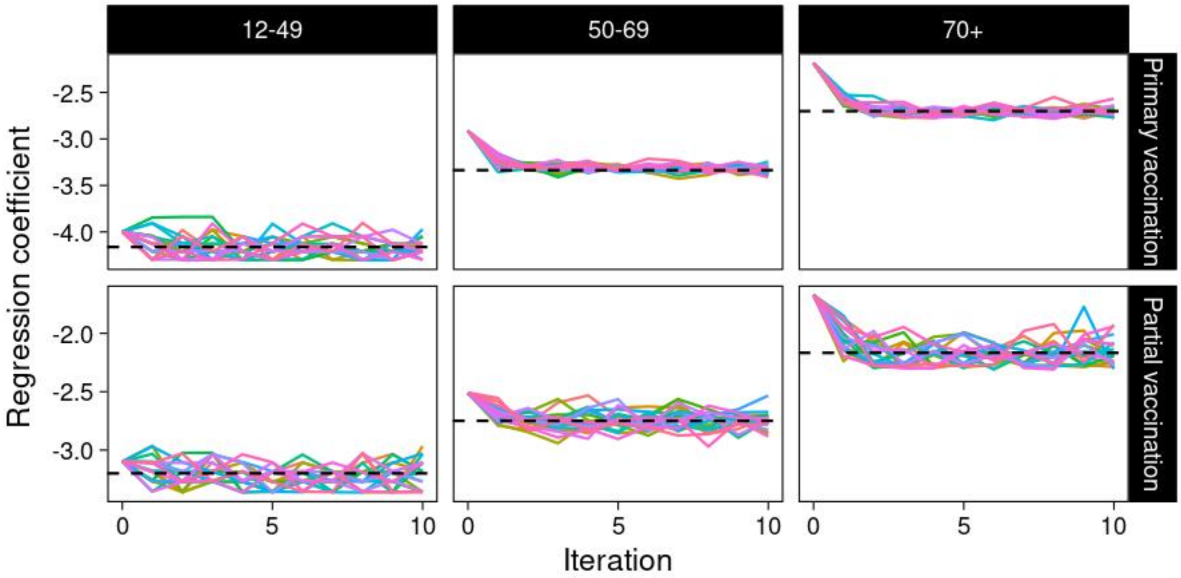

Figure S4: Uncorrected and corrected VE (A) and RR (B) estimates under different assumed relative risks of endpoint when not providing consent

Abbreviations: RR: relative risk of the outcome for the non-consenting vaccinated individuals compared to the consenting vaccinated individuals; VE: vaccine effectiveness; Hosp: hospital; ICU: intensive care unit.

**A.**

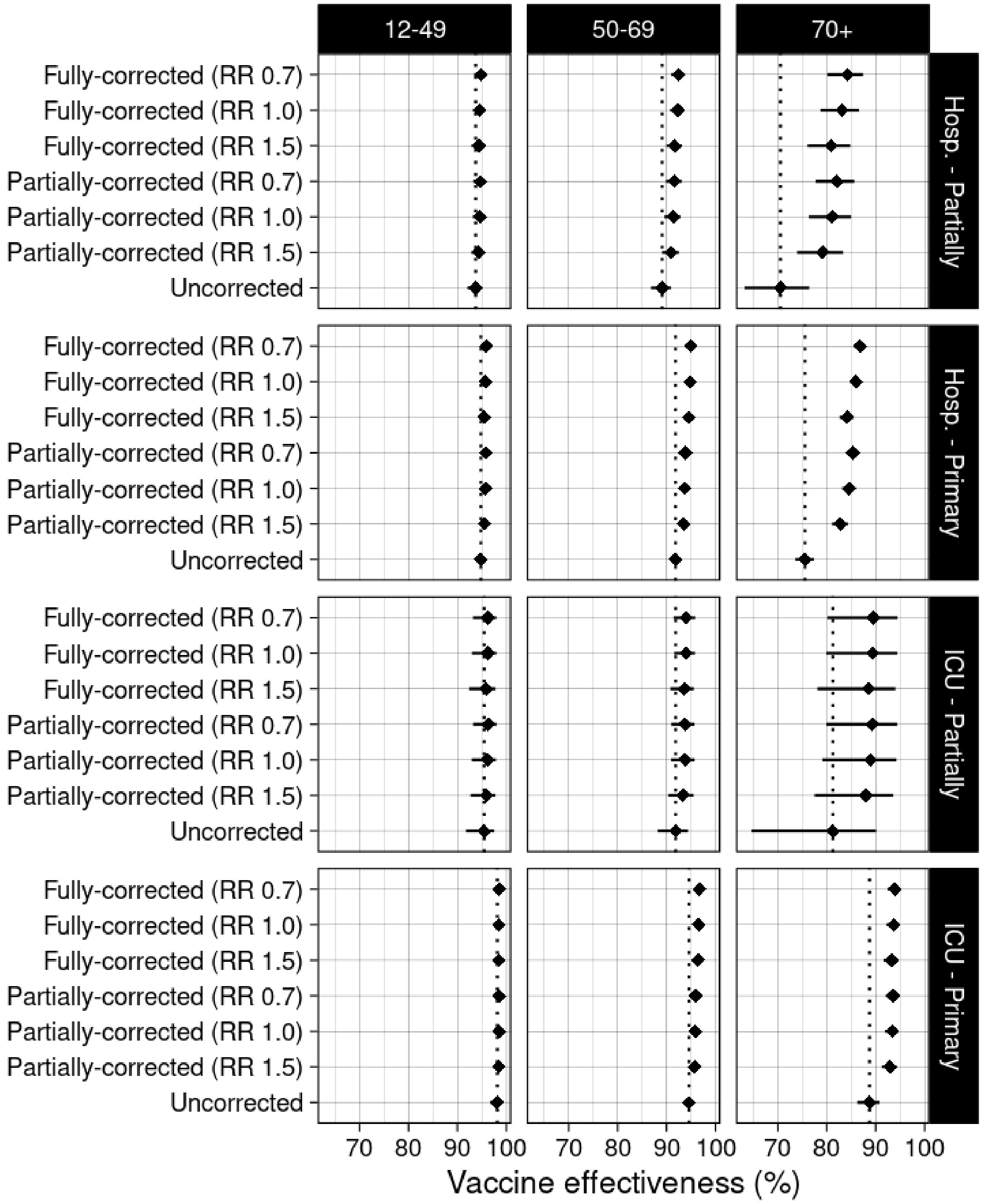

B.

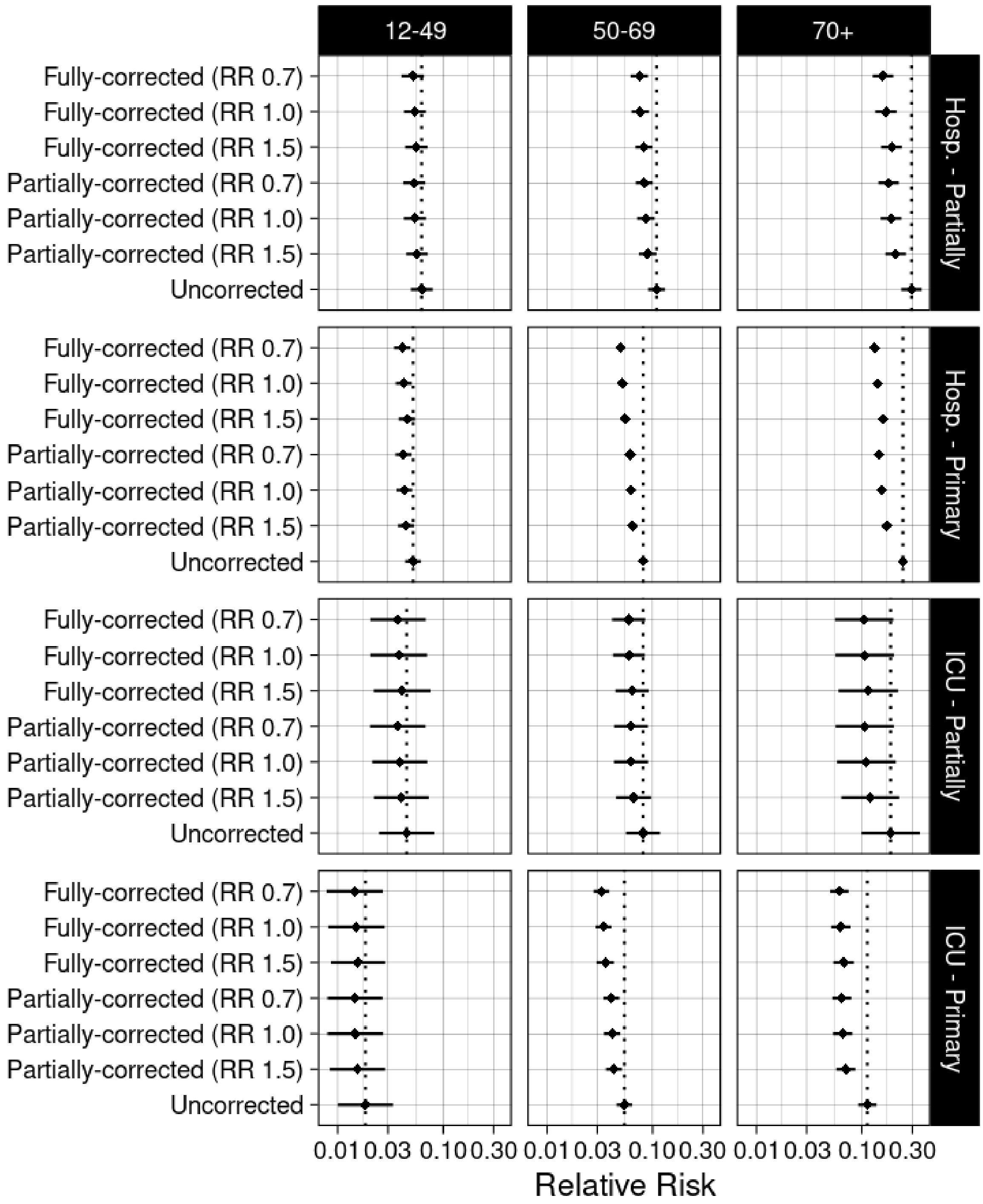
12-49
50-69
70+
Hosp. - Partially
Hosp. - Primary
ICU - Partially
ICU - Primary
Fully-corrected (RR 0.7)
Fully-corrected (RR 1.0)
Fully-corrected (RR 1.5)
Partially-corrected (RR 0.7)
Partially-corrected (RR 1.0)
Partially-corrected (RR 1.5)
Uncorrected
0.01 0.03 0.10 0.30
Relative Risk

Table S1: Uncorrected and corrected VE estimates assuming lower or higher risk of endpoint when not providing consent

| Vaccination status | Person years | Events | VE | VE bias | RR | RR bias |
|---|---|---|---|---|---|---|
| Hospitalization \| 12-49 years \| Uncorrected | | | | | | |
| Unvaccinated | 1,165,490 | 1,989 | - | - | - | - |
| Partially vaccinated | 511,966 | 71 | 93.8% (92.1; 95.1) | - | 0.062 (0.049-0.079) | - |
| Primary vaccinated | 1,176,241 | 143 | 94.8% (93.8; 95.6) | - | 0.052 (0.044-0.062) | - |
| Hospitalization \| 12-49 years \| Partially-corrected (RR 1.5) | | | | | | |
| Unvaccinated | 1,039,598 | 1,964 | - | - | - | - |
| Partially vaccinated | 553,464 | 80 | 94.3% (92.8; 95.5) | -0.6% | 0.057 (0.045-0.072) | 1.11 |
| Primary vaccinated | 1,260,635 | 158 | 95.6% (94.7; 96.3) | -0.7% | 0.044 (0.037-0.053) | 1.17 |
| Hospitalization \| 12-49 years \| Partially-corrected (RR 1.0) | | | | | | |
| Unvaccinated | 1,039,598 | 1,973 | - | - | - | - |
| Partially vaccinated | 553,464 | 76 | 94.6% (93.1; 95.8) | -0.9% | 0.054 (0.042-0.069) | 1.16 |
| Primary vaccinated | 1,260,635 | 152 | 95.7% (94.9; 96.4) | -0.9% | 0.043 (0.036-0.051) | 1.21 |
| Hospitalization \| 12-49 years \| Partially-corrected (RR 0.7) | | | | | | |
| Unvaccinated | 1,039,598 | 1,976 | - | - | - | - |
| Partially vaccinated | 553,464 | 75 | 94.7% (93.3; 95.8) | -1.0% | 0.053 (0.042-0.067) | 1.18 |
| Primary vaccinated | 1,260,635 | 150 | 95.8% (95.0; 96.5) | -1.0% | 0.042 (0.035-0.050) | 1.23 |
| Hospitalization \| 12-49 years \| Fully-corrected (RR 1.5) | | | | | | |
| Unvaccinated | 1,028,344 | 1,957 | - | - | - | - |
| Partially vaccinated | 554,978 | 80 | 94.4% (92.9; 95.6) | -0.7% | 0.056 (0.044-0.071) | 1.12 |
| Primary vaccinated | 1,270,376 | 164 | 95.5% (94.6; 96.2) | -0.7% | 0.045 (0.038-0.054) | 1.15 |
| Hospitalization \| 12-49 years \| Fully-corrected (RR 1.0) | | | | | | |
| Unvaccinated | 1,028,495 | 1,969 | - | - | - | - |

| Vaccination status | Person years | Events | VE | VE bias | RR | RR bias |
|---|---|---|---|---|---|---|
| Partially vaccinated | 554,660 | 78 | 94.6% (93.2; 95.8) | -0.9% | 0.054 (0.042-0.068) | 1.16 |
| Primary vaccinated | 1,270,542 | 155 | 95.8% (95.0; 96.5) | -1.0% | 0.042 (0.035-0.050) | 1.23 |
| Hospitalization \| 12-49 years \| Fully-corrected (RR 0.7) | | | | | | |
| Unvaccinated | 1,028,402 | 1,977 | - | - | - | - |
| Partially vaccinated | 554,936 | 75 | 94.9% (93.4; 96.0) | -1.1% | 0.051 (0.040-0.066) | 1.21 |
| Primary vaccinated | 1,270,359 | 150 | 95.9% (95.1; 96.6) | -1.1% | 0.041 (0.034-0.049) | 1.27 |
| Hospitalization \| 50-69 years \| Uncorrected | | | | | | |
| Unvaccinated | 279,752 | 1,978 | - | - | - | - |
| Partially vaccinated | 175,829 | 126 | 89.1% (86.8; 91.0) | - | 0.109 (0.090-0.132) | - |
| Primary vaccinated | 1,208,038 | 806 | 91.8% (91.1; 92.5) | - | 0.082 (0.075-0.089) | - |
| Hospitalization \| 50-69 years \| Partially-corrected (RR 1.5) | | | | | | |
| Unvaccinated | 208,944 | 1,912 | - | - | - | - |
| Partially vaccinated | 187,771 | 140 | 91.0% (89.1; 92.6) | -1.9% | 0.090 (0.074-0.109) | 1.21 |
| Primary vaccinated | 1,266,904 | 857 | 93.5% (92.9; 94.1) | -1.7% | 0.065 (0.059-0.071) | 1.26 |
| Hospitalization \| 50-69 years \| Partially-corrected (RR 1.0) | | | | | | |
| Unvaccinated | 208,944 | 1,934 | - | - | - | - |
| Partially vaccinated | 187,771 | 135 | 91.4% (89.6; 92.8) | -2.3% | 0.086 (0.072-0.104) | 1.26 |
| Primary vaccinated | 1,266,904 | 839 | 93.7% (93.1; 94.3) | -1.9% | 0.063 (0.057-0.069) | 1.30 |
| Hospitalization \| 50-69 years \| Partially-corrected (RR 0.7) | | | | | | |
| Unvaccinated | 208,944 | 1,949 | - | - | - | - |
| Partially vaccinated | 187,771 | 132 | 91.7% (89.9; 93.1) | -2.6% | 0.083 (0.069-0.101) | 1.31 |
| Primary vaccinated | 1,266,904 | 828 | 93.9% (93.3; 94.4) | -2.0% | 0.061 (0.056-0.067) | 1.33 |
| Hospitalization \| 50-69 years \| Fully-corrected (RR 1.5) | | | | | | |

| Vaccination status | Person years | Events | VE | VE bias | RR | RR bias |
|---|---|---|---|---|---|---|
| Unvaccinated | 178,082 | 1,839 | - | - | - | - |
| Partially vaccinated | 190,142 | 155 | 91.7% (90.0; 93.1) | -2.6% | 0.083 (0.069-0.100) | 1.32 |
| Primary vaccinated | 1,295,394 | 915 | 94.5% (94.0; 95.0) | -2.7% | 0.055 (0.050-0.060) | 1.49 |
| Hospitalization \| 50-69 years \| Fully-corrected (RR 1.0) | | | | | | |
| Unvaccinated | 178,116 | 1,878 | - | - | - | - |
| Partially vaccinated | 189,874 | 146 | 92.3% (90.8; 93.7) | -3.3% | 0.077 (0.063-0.092) | 1.43 |
| Primary vaccinated | 1,295,628 | 885 | 94.8% (94.3; 95.3) | -3.0% | 0.052 (0.047-0.057) | 1.58 |
| Hospitalization \| 50-69 years \| Fully-corrected (RR 0.7) | | | | | | |
| Unvaccinated | 178,138 | 1,899 | - | - | - | - |
| Partially vaccinated | 190,059 | 144 | 92.5% (91.0; 93.8) | -3.4% | 0.075 (0.062-0.090) | 1.46 |
| Primary vaccinated | 1,295,421 | 865 | 95.0% (94.5; 95.4) | -3.2% | 0.050 (0.046-0.055) | 1.63 |
| Hospitalization \| 70+ years \| Uncorrected | | | | | | |
| Unvaccinated | 123,945 | 1,432 | - | - | - | - |
| Partially vaccinated | 27,281 | 86 | 70.5% (63.2; 76.4) | - | 0.295 (0.236-0.368) | - |
| Primary vaccinated | 764,788 | 2,173 | 75.5% (73.5; 77.4) | - | 0.245 (0.226-0.265) | - |
| Hospitalization \| 70+ years \| Partially-corrected (RR 1.5) | | | | | | |
| Unvaccinated | 71,075 | 1,193 | - | - | - | - |
| Partially vaccinated | 28,820 | 93 | 79.1% (73.9; 83.3) | -8.6% | 0.209 (0.167-0.261) | 1.41 |
| Primary vaccinated | 816,120 | 2,404 | 82.8% (81.1; 84.4) | -7.3% | 0.172 (0.156-0.189) | 1.42 |
| Hospitalization \| 70+ years \| Partially-corrected (RR 1.0) | | | | | | |
| Unvaccinated | 71,075 | 1,279 | - | - | - | - |
| Partially vaccinated | 28,820 | 91 | 81.1% (76.3; 85.0) | -10.6% | 0.189 (0.150-0.237) | 1.56 |
| Primary vaccinated | 816,120 | 2,320 | 84.5% (83.1; 85.8) | -9.0% | 0.155 (0.142-0.169) | 1.58 |

| Vaccination status | Person years | Events | VE | VE bias | RR | RR bias |
|---|---|---|---|---|---|---|
| Hospitalization \| 70+ years \| Partially-corrected (RR 0.7) | | | | | | |
| Unvaccinated | 71,075 | 1,326 | - | - | - | - |
| Partially vaccinated | 28,820 | 89 | 82.1% (77.7; 85.7) | -11.6% | 0.179 (0.143-0.223) | 1.65 |
| Primary vaccinated | 816,120 | 2,275 | 85.4% (84.1; 86.6) | -9.8% | 0.146 (0.134-0.159) | 1.67 |
| Hospitalization \| 70+ years \| Fully-corrected (RR 1.5) | | | | | | |
| Unvaccinated | 65,063 | 1,170 | - | - | - | - |
| Partially vaccinated | 29,625 | 97 | 80.9% (76.1; 84.8) | -10.4% | 0.191 (0.152-0.239) | 1.54 |
| Primary vaccinated | 821,326 | 2,422 | 84.1% (82.6; 85.5) | -8.6% | 0.159 (0.145-0.174) | 1.54 |
| Hospitalization \| 70+ years \| Fully-corrected (RR 1.0) | | | | | | |
| Unvaccinated | 65,074 | 1,265 | - | - | - | - |
| Partially vaccinated | 29,506 | 93 | 83.1% (78.7; 86.6) | -12.6% | 0.169 (0.134-0.213) | 1.75 |
| Primary vaccinated | 821,435 | 2,331 | 85.9% (84.7; 87.1) | -10.4% | 0.141 (0.129-0.153) | 1.74 |
| Hospitalization \| 70+ years \| Fully-corrected (RR 0.7) | | | | | | |
| Unvaccinated | 65,093 | 1,318 | - | - | - | - |
| Partially vaccinated | 29,607 | 91 | 84.2% (80.2; 87.4) | -13.7% | 0.158 (0.126-0.198) | 1.86 |
| Primary vaccinated | 821,315 | 2,280 | 86.8% (85.6; 87.8) | -11.2% | 0.132 (0.122-0.144) | 1.85 |
| ICU admission \| 12-49 years \| Uncorrected | | | | | | |
| Unvaccinated | 1,165,490 | 400 | - | - | - | - |
| Partially vaccinated | 511,966 | 11 | 95.5% (91.8; 97.5) | - | 0.045 (0.025-0.082) | - |
| Primary vaccinated | 1,176,241 | 11 | 98.2% (96.6; 99.0) | - | 0.018 (0.010-0.034) | - |
| ICU admission \| 12-49 years \| Partially-corrected (RR 1.5) | | | | | | |
| Unvaccinated | 1,039,598 | 397 | - | - | - | - |
| Partially vaccinated | 553,464 | 12 | 96.0% (92.7; 97.8) | -0.5% | 0.040 (0.022-0.073) | 1.12 |

| Vaccination status | Person years | Events | VE | VE bias | RR | RR bias |
|---|---|---|---|---|---|---|
| Primary vaccinated | 1,260,635 | 12 | 98.5% (97.2; 99.2) | -0.3% | 0.015 (0.008-0.028) | 1.19 |
| ICU admission \| 12-49 years \| Partially-corrected (RR 1.0) | | | | | | |
| Unvaccinated | 1,039,598 | 398 | - | - | - | - |
| Partially vaccinated | 553,464 | 12 | 96.1% (92.9; 97.9) | -0.6% | 0.039 (0.021-0.071) | 1.16 |
| Primary vaccinated | 1,260,635 | 11 | 98.5% (97.3; 99.2) | -0.4% | 0.015 (0.008-0.027) | 1.25 |
| ICU admission \| 12-49 years \| Partially-corrected (RR 0.7) | | | | | | |
| Unvaccinated | 1,039,598 | 399 | - | - | - | - |
| Partially vaccinated | 553,464 | 11 | 96.3% (93.2; 98.0) | -0.8% | 0.037 (0.020-0.068) | 1.22 |
| Primary vaccinated | 1,260,635 | 11 | 98.5% (97.3; 99.2) | -0.4% | 0.015 (0.008-0.027) | 1.26 |
| ICU admission \| 12-49 years \| Fully-corrected (RR 1.5) | | | | | | |
| Unvaccinated | 1,028,467 | 396 | - | - | - | - |
| Partially vaccinated | 554,760 | 12 | 95.9% (92.4; 97.8) | -0.4% | 0.041 (0.022-0.076) | 1.11 |
| Primary vaccinated | 1,270,470 | 12 | 98.4% (97.2; 99.1) | -0.3% | 0.016 (0.009-0.028) | 1.18 |
| ICU admission \| 12-49 years \| Fully-corrected (RR 1.0) | | | | | | |
| Unvaccinated | 1,028,519 | 397 | - | - | - | - |
| Partially vaccinated | 554,615 | 12 | 96.2% (93.0; 98.0) | -0.7% | 0.038 (0.020-0.070) | 1.19 |
| Primary vaccinated | 1,270,563 | 12 | 98.5% (97.2; 99.2) | -0.3% | 0.015 (0.008-0.028) | 1.22 |
| ICU admission \| 12-49 years \| Fully-corrected (RR 0.7) | | | | | | |
| Unvaccinated | 1,028,468 | 398 | - | - | - | - |
| Partially vaccinated | 554,809 | 11 | 96.3% (93.2; 98.0) | -0.8% | 0.037 (0.020-0.068) | 1.21 |
| Primary vaccinated | 1,270,420 | 11 | 98.5% (97.3; 99.2) | -0.4% | 0.015 (0.008-0.027) | 1.26 |
| ICU admission \| 50-69 years \| Uncorrected | | | | | | |
| Unvaccinated | 279,752 | 634 | - | - | - | - |

| Vaccination status | Person years | Events | VE | VE bias | RR | RR bias |
|---|---|---|---|---|---|---|
| Partially vaccinated | 175,829 | 30 | 91.9% (88.2; 94.4) | - | 0.081 (0.056-0.118) | - |
| Primary vaccinated | 1,208,038 | 170 | 94.6% (93.6; 95.4) | - | 0.054 (0.046-0.064) | - |
| ICU admission \| 50-69 years \| Partially-corrected (RR 1.5) | | | | | | |
| Unvaccinated | 208,944 | 619 | - | - | - | - |
| Partially vaccinated | 187,771 | 32 | 93.4% (90.3; 95.5) | -1.5% | 0.066 (0.045-0.097) | 1.23 |
| Primary vaccinated | 1,266,904 | 181 | 95.7% (94.9; 96.4) | -1.1% | 0.043 (0.036-0.051) | 1.26 |
| ICU admission \| 50-69 years \| Partially-corrected (RR 1.0) | | | | | | |
| Unvaccinated | 208,944 | 625 | - | - | - | - |
| Partially vaccinated | 187,771 | 31 | 93.8% (90.9; 95.7) | -1.9% | 0.062 (0.043-0.091) | 1.31 |
| Primary vaccinated | 1,266,904 | 176 | 95.9% (95.1; 96.5) | -1.3% | 0.041 (0.035-0.049) | 1.31 |
| ICU admission \| 50-69 years \| Partially-corrected (RR 0.7) | | | | | | |
| Unvaccinated | 208,944 | 627 | - | - | - | - |
| Partially vaccinated | 187,771 | 31 | 93.8% (91.0; 95.7) | -1.9% | 0.062 (0.043-0.090) | 1.31 |
| Primary vaccinated | 1,266,904 | 174 | 95.9% (95.2; 96.6) | -1.3% | 0.041 (0.034-0.048) | 1.33 |
| ICU admission \| 50-69 years \| Fully-corrected (RR 1.5) | | | | | | |
| Unvaccinated | 178,176 | 599 | - | - | - | - |
| Partially vaccinated | 189,906 | 39 | 93.6% (90.8; 95.5) | -1.7% | 0.064 (0.045-0.092) | 1.27 |
| Primary vaccinated | 1,295,536 | 195 | 96.4% (95.7; 97.0) | -1.9% | 0.036 (0.030-0.043) | 1.52 |
| ICU admission \| 50-69 years \| Fully-corrected (RR 1.0) | | | | | | |
| Unvaccinated | 178,146 | 605 | - | - | - | - |
| Partially vaccinated | 189,794 | 36 | 94.0% (91.5; 95.8) | -2.2% | 0.060 (0.042-0.085) | 1.36 |
| Primary vaccinated | 1,295,679 | 191 | 96.6% (95.9; 97.1) | -2.0% | 0.034 (0.029-0.041) | 1.58 |
| ICU admission \| 50-69 years \| Fully-corrected (RR 0.7) | | | | | | |

| Vaccination status | Person years | Events | VE | VE bias | RR | RR bias |
|---|---|---|---|---|---|---|
| Unvaccinated | 178,237 | 612 | - | - | - | - |
| Partially vaccinated | 189,930 | 37 | 94.0% (91.4; 95.9) | -2.2% | 0.060 (0.041-0.086) | 1.37 |
| Primary vaccinated | 1,295,451 | 184 | 96.7% (96.1; 97.2) | -2.1% | 0.033 (0.028-0.039) | 1.65 |
| ICU admission \| 70+ years \| Uncorrected | | | | | | |
| Unvaccinated | 123,945 | 244 | - | - | - | - |
| Partially vaccinated | 27,281 | 10 | 81.3% (64.6; 90.1) | - | 0.187 (0.099-0.354) | - |
| Primary vaccinated | 764,788 | 182 | 88.7% (86.2; 90.8) | - | 0.113 (0.092-0.138) | - |
| ICU admission \| 70+ years \| Partially-corrected (RR 1.5) | | | | | | |
| Unvaccinated | 71,075 | 226 | - | - | - | - |
| Partially vaccinated | 28,820 | 11 | 88.0% (77.5; 93.6) | -6.7% | 0.120 (0.064-0.225) | 1.56 |
| Primary vaccinated | 816,120 | 198 | 92.9% (91.3; 94.3) | -4.2% | 0.071 (0.057-0.087) | 1.59 |
| ICU admission \| 70+ years \| Partially-corrected (RR 1.0) | | | | | | |
| Unvaccinated | 71,075 | 234 | - | - | - | - |
| Partially vaccinated | 28,820 | 10 | 89.0% (79.1; 94.2) | -7.7% | 0.110 (0.058-0.209) | 1.70 |
| Primary vaccinated | 816,120 | 190 | 93.4% (91.9; 94.7) | -4.7% | 0.066 (0.053-0.081) | 1.71 |
| ICU admission \| 70+ years \| Partially-corrected (RR 0.7) | | | | | | |
| Unvaccinated | 71,075 | 236 | - | - | - | - |
| Partially vaccinated | 28,820 | 10 | 89.4% (79.9; 94.4) | -8.1% | 0.106 (0.056-0.201) | 1.76 |
| Primary vaccinated | 816,120 | 189 | 93.5% (92.1; 94.7) | -4.8% | 0.065 (0.053-0.079) | 1.74 |
| ICU admission \| 70+ years \| Fully-corrected (RR 1.5) | | | | | | |
| Unvaccinated | 65,091 | 226 | - | - | - | - |
| Partially vaccinated | 29,539 | 11 | 88.6% (78.1; 94.0) | -7.3% | 0.114 (0.060-0.219) | 1.64 |
| Primary vaccinated | 821,384 | 198 | 93.3% (91.6; 94.6) | -4.5% | 0.067 (0.054-0.084) | 1.68 |

| Vaccination status | Person years | Events | VE | VE bias | RR | RR bias |
|---|---|---|---|---|---|---|
| ICU admission \| 70+ years \| Fully-corrected (RR 1.0) | | | | | | |
| Unvaccinated | 65,117 | 232 | - | - | - | - |
| Partially vaccinated | 29,443 | 10 | 89.4% (79.9; 94.4) | -8.1% | 0.106 (0.056-0.201) | 1.76 |
| Primary vaccinated | 821,454 | 192 | 93.7% (92.2; 94.9) | -5.0% | 0.063 (0.051-0.078) | 1.79 |
| ICU admission \| 70+ years \| Fully-corrected (RR 0.7) | | | | | | |
| Unvaccinated | 65,102 | 235 | - | - | - | - |
| Partially vaccinated | 29,546 | 10 | 89.5% (80.2; 94.4) | -8.2% | 0.105 (0.056-0.198) | 1.78 |
| Primary vaccinated | 821,368 | 189 | 93.9% (92.5; 95.0) | -5.1% | 0.061 (0.050-0.075) | 1.84 |

Abbreviations: VE: vaccine effectiveness, RR: relative risk of the outcome for the non-consenting vaccinated individuals compared to the consenting vaccinated individuals.

#### Table S2: Uncorrected and corrected regression coefficients, standard errors and fraction of missing information

For models assuming incidence of the endpoint independent of providing consent (RR 1.0).

| | Partially vaccinated | | | Primary vaccinated | | |
|---|---|---|---|---|---|---|
| **Model** | **Coefficient** | **SE** | **$y_{mis}$[1]** | **Coefficient** | **SE** | **$y_{mis}$[1]** |
| Hospitalization \| 12-49 years | | | | | | |
| Uncorrected | -2.773 | 0.122 | - | -2.960 | 0.088 | - |
| Partially-corrected | -2.923 | 0.125 | 0.10 (0.06-0.17) | -3.152 | 0.088 | 0.05 (0.03-0.09) |
| Fully-corrected | -2.922 | 0.122 | 0.09 (0.05-0.15) | -3.165 | 0.089 | 0.08 (0.04-0.13) |
| Hospitalization \| 50-69 years | | | | | | |
| Uncorrected | -2.216 | 0.096 | - | -2.506 | 0.046 | - |
| Partially-corrected | -2.449 | 0.096 | 0.08 (0.04-0.13) | -2.768 | 0.047 | 0.05 (0.03-0.09) |
| Fully-corrected | -2.570 | 0.096 | 0.14 (0.08-0.23) | -2.962 | 0.048 | 0.11 (0.06-0.19) |
| Hospitalization \| 70+ years | | | | | | |
| Uncorrected | -1.222 | 0.113 | - | -1.408 | 0.041 | - |
| Partially-corrected | -1.667 | 0.116 | 0.10 (0.05-0.17) | -1.867 | 0.045 | 0.17 (0.10-0.27) |
| Fully-corrected | -1.779 | 0.117 | 0.14 (0.08-0.23) | -1.961 | 0.043 | 0.16 (0.09-0.26) |
| ICU admission \| 12-49 years | | | | | | |
| Uncorrected | -3.098 | 0.308 | - | -3.998 | 0.307 | - |
| Partially-corrected | -3.249 | 0.307 | 0.08 (0.04-0.13) | -4.223 | 0.309 | 0.05 (0.03-0.10) |
| Fully-corrected | -3.274 | 0.317 | 0.13 (0.07-0.21) | -4.199 | 0.315 | 0.13 (0.07-0.21) |
| ICU admission \| 50-69 years | | | | | | |
| Uncorrected | -2.509 | 0.189 | - | -2.916 | 0.087 | - |
| Partially-corrected | -2.776 | 0.191 | 0.06 (0.03-0.10) | -3.186 | 0.089 | 0.07 (0.04-0.12) |
| Fully-corrected | -2.820 | 0.178 | 0.07 (0.04-0.13) | -3.371 | 0.089 | 0.11 (0.06-0.18) |
| ICU admission \| 70+ years | | | | | | |
| Uncorrected | -1.674 | 0.324 | - | -2.184 | 0.103 | - |
| Partially-corrected | -2.205 | 0.328 | 0.06 (0.03-0.10) | -2.722 | 0.107 | 0.09 (0.05-0.16) |
| Fully-corrected | -2.242 | 0.325 | 0.05 (0.03-0.09) | -2.766 | 0.108 | 0.10 (0.05-0.16) |

[1]$y_{mis}$ is the fraction of information that is missing about a model parameter. Numbers between brackets represent the 95% confidence interval. Statistical efficiency of the imputation model, compared to having consent in 100% of vaccinated individuals, is given as (1 - $y_{mis}$) * 100%.
Abbreviations: RR: relative risk; SE: standard error.

### References

1. van Iersel S, McDonald SA, de Gier B, Knol MJ, de Melker HE, Henri van Werkhoven CH, et al. Number of COVID-19 hospitalisations averted by vaccination: Estimates for the Netherlands, January 6, 2021 through August 30, 2022. Vaccine. 2023;41(26):3847-54.
2. Gier Bd, Kooijman M, Kemmeren J, Keizer Nd, Dongelmans D, Iersel SCJLv, et al. COVID-19 vaccine effectiveness against hospitalizations and ICU admissions in the Netherlands, April- August 2021. medRxiv. 2021:2021.09.15.21263613.
3. Shao W, Chen X, Zheng C, Liu H, Wang G, Zhang B, et al. Effectiveness of COVID-19 vaccines against SARS-CoV-2 variants of concern in real-world: a literature review and meta-analysis. Emerg Microbes Infect. 2022;11(1):2383-92.
4. Rubin DB. Multiple Imputation for Nonresponse in Surveys. New York: Wiley; 1987.
5. von Hippel PT. How Many Imputations Do You Need? A Two-stage Calculation Using a Quadratic Rule. Sociological Methods & Research. 2020;49(3):699-718.
